# A Proteome-wide, Multi-Omics Analysis Implicates Novel Protein Dysregulation in Post-Traumatic Stress Disorder

**DOI:** 10.1101/2023.05.05.23289589

**Authors:** Jiawei Wang, Hongyu Li, Rashaun Wilson, Weiwei Wang, TuKiet T. Lam, Traumatic Stress Brain Research Group, David A. Lewis, Jill Glausier, Paul E. Holtzheimer, Matthew J. Friedman, Kenneth R. Williams, Marina R. Picciotto, Angus C. Nairn, John H. Krystal, Ronald S. Duman, Hongyu Zhao, Matthew J. Girgenti

**Author notes:** Correspondence (M.J.G.). Deceased (Ronald S. Duman).

## Abstract

Post-traumatic stress disorder (PTSD) is a common and disabling psychiatric disorder. Here we present findings from the first proteome-wide study of the postmortem PTSD brain. We performed tandem mass spectrometry on large cohort of donors (N = 66) in two prefrontal cortical areas and found differentially expressed proteins and co-expression modules disturbed in PTSD. Integrative analysis pointed to *hsa-mir-589* as a regulatory miRNA responsible for disruptions in neuronal protein networks for PTSD, including the GABA vesicular transporter, SLC32A1. In addition, we identified significant enrichment of risk genes for Alzheimer’s Disease (N= 94,403), major depression (N = 807,553), and schizophrenia (N = 35,802) within PTSD co-expression protein modules, suggesting shared molecular pathology. Our findings highlight the altered proteomic landscape of postmortem PTSD brain and provide a novel framework for future studies integrating proteomic profiling with transcriptomics in postmortem human brain tissue.

## Introduction

Post-traumatic stress disorder (PTSD) is a severe mental illness that affects millions of people worldwide^1,2^. Patients with PTSD typically have symptoms that include re-experiencing of traumatic memories^3,4^, hyperarousal, emotional numbing, dysphoric mood, and avoidance. In addition, PTSD is frequently comorbid with other psychiatric disorders, such as major depressive disorder, which occurs in 51-82% of PTSD cases^5–7^. PTSD heritability estimates range from 30-40%^8–10^ and recent evidence suggests PTSD is highly polygenic^8,11,12^. To date, only a few genomic loci have been implicated in the risk for PTSD. Recent large meta-analyses by the Million Veteran Program and the Psychiatric Genomics Consortium revealed three significant loci and 15 risk loci for quantitative symptom traits^8,11–13^. Transcriptomic studies using human postmortem prefrontal cortical (PFC) tissue have linked dysregulation of biological processes to PTSD, including GABA signaling, inflammation and cytokine effects, and glucocorticoid signaling^14–16^.

Tandem mass spectrometry (MS/MS) has become an indispensable tool for obtaining unbiased, high resolution proteomic data. Whole-proteome analysis is essential for understanding the molecular facets of the human brain, because proteins and their changes provide unique insight into the state of the cell. The entire neuroproteome can only be profiled using mass spectrometry, which has comparable throughput and resolution of other functional genomic techniques. To fully understand the functional and system-level roles of CNS cells in disease, quantitative investigation of the thousands of proteins expressed in each of the likely hundreds of different neuronal and non-neuronal cell types is essential. To better understand the dynamics of RNA and protein expression, the roles of post-transcriptional modifications, and the trafficking of transcripts and proteins must be integrated with other functional genomic data. Despite the potential value, few studies have been conducted in human postmortem brains of donors with major psychiatric illness. And fewer still have sought to integrate this data with other genomic modalities. Additionally, as the final step of the central dogma of molecular biology, protein sequence and abundance may be the most relevant genomic level to identify potential therapeutic points of intervention.

Systems-level analyses of multi-omics datasets are essential tools for identifying molecular targets for disease processes beyond what coding gene expression alone could reveal^17^. To our knowledge, no postmortem genomics studies have been conducted matching mRNA, miRNA, and proteomics from cases and controls from the same postmortem human brain donors. To identify potential causal brain proteins in PTSD, we endeavored to generate a first of its kind dataset. We performed proteomic tandem mass spectrometry of postmortem brains from individuals with PTSD, a psychiatric comparison group (MDD), and neurotypical controls and coupled this with genome-wide expression profiling of mRNA and miRNAs. We examined tissue from two prefrontal cortical regions: dorsolateral prefrontal cortex (DLPFC, Brodmann area 9/46) and subgenual prefrontal cortex (sgPFC, Brodmann area 25). These two regions were chosen based on previous clinical evidence of functional engagement in PTSD^18,19^. We developed a multi-modal bioinformatics analysis pipeline to link perturbations in proteomic alteration to miRNA expression and transcriptomic changes in the PTSD-affected brain.

We identified PTSD-specific protein differential expression signatures and co-expression patterns, including down-regulation of interneuron-specific modules containing GABAergic proteins SLC32A1 and NEGR1. Brain proteome-wide association analysis identified significant enrichment of risk genes for schizophrenia, major depression, and Alzheimer’s disease within PTSD proteomic networks, pointing to shared molecular pathology and risk. In addition, we found that numerous miRNAs were upregulated in the PTSD brain, including *hsa-mir-589* and *hsa-mir-6786* and integrative analysis identified miRNAs enriched for disease-associated proteins and protein modules. Our findings highlight the use of large-scale multi-omics systems biology to unravel the effects of the neuroproteome in psychiatric disorders.

## Results

### Differentially expressed proteomic signatures in PTSD and integration with gene expression

We analyzed postmortem brain samples from a large cohort (N=66), including neurotypical control subjects (CON, N=22), subjects with major depressive disorder (MDD, N=22), and subjects with posttraumatic stress disorder (PTSD, N=22). We profiled 122 postmortem brain tissue samples from PTSD and MDD cases and controls (CON) across 2 cortical regions: DLPFC (Broadmann area 9) and sgPFC (Broadmann area 25) (**Fig. 1A**, **Table 1**). We performed tandem mass spectrometry (LC MS/MS) on the samples to measure protein abundance levels using data-independent acquisition (DIA)^20^ (**Fig. 1A**). After extensive and rigorous quality control of LC MS/MS (see Methods), we performed differential expression analysis for 109 samples from 57 unique donors. Detailed sample information with corresponding demographics are listed in **Table 1**. We identified 2,775 proteins initially and 2,598 after removing those with missing values. Proteomic coverage tended to overlap with the transcripts with the highest expression levels (**Fig. 1B**). Principal Component Analysis (PCA) was performed to assess the effects of demographic confounders in protein expression variation. Regional differences in proteomic profiles were captured by PC1 (accounting for 29% of the variance) (**Extended Data Figure 1A**). Previous characterization of the transcriptome of these donors identified significant differences between males and females^14^. Surprisingly, sex accounted for less of the variance in this dataset (**Extended Data Figure 1A&B**).

**Figure 1.**
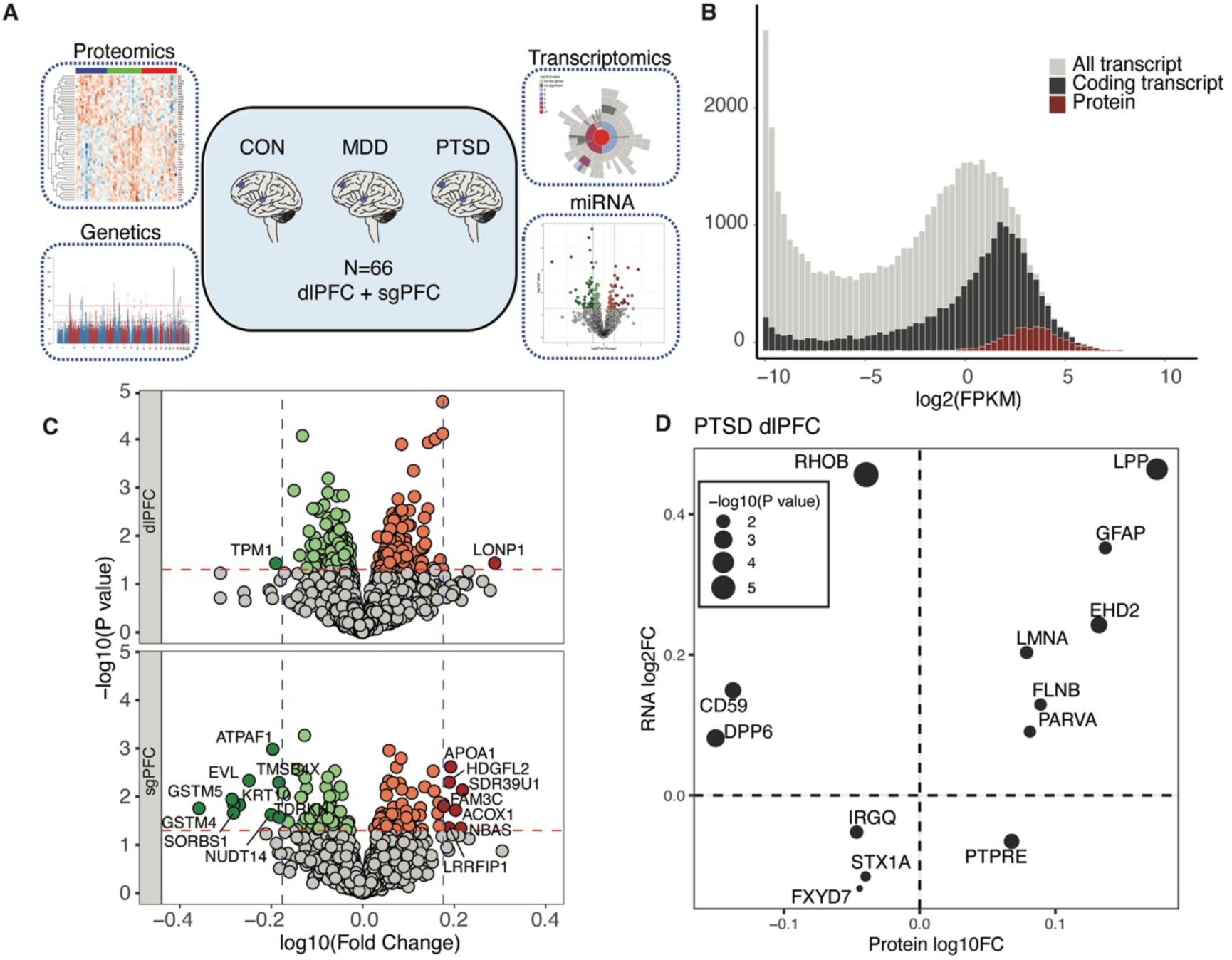
Genomic data overview and differential expression analysis of proteins in PTSD. (**A**) A schematic overview of the study design and multiple -omics datasets employed in this study. (**B**) Distribution of gene expression (all genes; gray), peptides (red) correspond to the majority of higher abundance mRNAs (RNA coding genes, black). (**C**) Volcano plots showing DEPs in DLPFC (top) and sgPFC (bottom) of PTSD brains. Orange (*P* < 0.05) and red (log fold change >0.18 and *P* < 0.05) indicate upregulated proteins and green (*P* < 0.05) and dark green (log fold change <-0.18) and *P* < 0.05) indicate down regulated proteins. (**D**) Correlation between transcript and protein differential expression in DLPFC of PTSD brains (Adjusted *R*^2^ = 0.12, robust regression *P =* 0.067).

**Table 1.** Demographics of Sample donors.

|  | **Control** | **MDD** | **PTSD** |
| --- | --- | --- | --- |
| N | 22 | 22 | 22 |
| RIN: dlPFC | 6.9 ± 1.2 | 7.4 ± 1.3 | 7.7 ± 0.9 |
| RIN: sgPFC | 8.1 ± 1.0 | 8.2 ± 0.8 | 8.2 ± 1.1 |
| Sex (%Male) | 50 | 50 | 50 |
| Race (%White) | 77.3 | 81.8 | 81.8 |
| Age at death | 47.8 ± 11.7 | 45.9 ± 12.34 | 46.0 ± 11.4 |
| Postmortem interval (PMI) | 18.8 ± 6.8 | 17.0 ± 5.6 | 16.0 ± 5.6 |
| Comorbid depression (%) | 0 | 100 | 50 |
| Tabaco (%ATOD) | 27.2 | 22.7 | 63.6 |
| Alcohol (%ATOD) | 0 | 16.7 | 43.9 |
| Opioids (%ATOD) | 0 | 68.2 | 72.7 |
| Antidepressants (%ATOD) | 0 | 72.7 | 68.2 |
| Manner of death (%Suicide) | 0 | 22.7 | 9.1 |

Differential expression analysis identified disease- and brain region-specific proteins (*P*-value < 0.05, **Fig. 1C**). Overlap of differentially expressed proteins (DEPs) was moderate between MDD and PTSD from the same brain regions (28% DLPFC, 10.7% sgPFC) but was generally low between different brain regions (5.3% and 2.8%) (**Extended Data Figure 2A**), indicating significant regional differences in the proteomes between DLPFC and sgPFC. Rank-rank hypergeometric overlap (RRHO) showed convergent proteomic changes between MDD and PTSD from the same brain region but not across regions within the same diagnostic cohort (**Extended Data Figure 2B**). In DLPFC, one protein, MACD1, passed the FDR-corrected threshold for PTSD (FDR= 0.043). Eight proteins were significantly changed in MDD after controlling for multiple comparisons (FDR < 0.05, **Supp Table 1**), including GNB4, RAC1, CNTFR, LY6H, CNTN1, DPYSL4, LAP3, and SLC17A7, and LAP3. No protein was found to be significantly changed in MDD or PTSD in sgPFC at an FDR < 0.05 cut-off. We next performed a secondary analysis within each of the two subregions to identify additional DEPs associated with PTSD and MDD diagnosis. Using a more liberal cutoff of *P* < 0.05, we identified 351 unique DEPs (DLPFC: 249, sgPFC: 112, 10 overlapping proteins) for PTSD. In MDD, we identified 361 unique DEPs (DLPFC: 243, sgPFC: 137, 19 proteins overlapping). A list of all DEPs and statistics across models appears in **Supp Table 1.**

**Figure 2.**
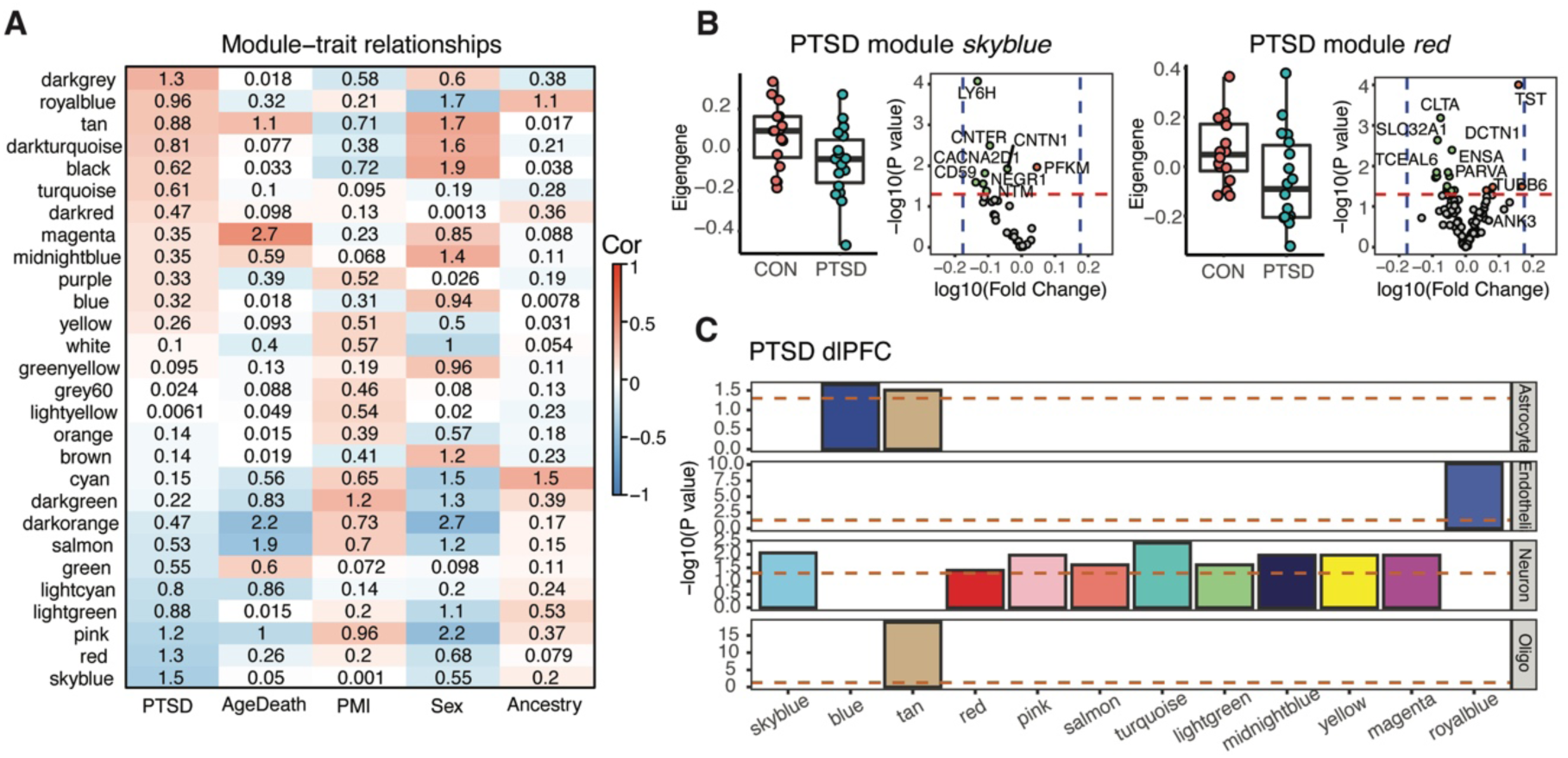
Network Co-expression analysis of proteomics data. (**A**) Module-trait correlation between protein expression correlations in the DLPFC of PTSD brains and demographic features (PrimaryDx, age of death, PMI, sex, ancestry). Color in each cell reflects to correlation between module eigenprotein and PrimaryDx, while the number represents -log_10_(*P*-value) of that correlation. (**B**) Boxplots of eigenproteins and volcano plots of the differential expression patterns of the proteins in PTSD-associated modules *skyblue* (left) and *red* (right) (vertical dash lines indicate log_10_(fold change) = ± 0.18). Module *skyblue*, *P* = 0.03; module *red*, *P* = 0.04. *P* indicates eigenprotein change significance between PTSD and CON(**C**) Cell type enrichment analysis showing the enrichment of cell type markers in protein modules. Brown dashed line indicates significantly enriched markers, *P* < 0.05.

We performed mRNA-seq on the same brain regions for this cohort of donors to understand the relationship between the PTSD transcriptome and proteome (**Fig. 1D**). In DLPFC, most PTSD-(**Fig. 1D**) and MDD-DEGs (**Extended Data Figure 2C**) exhibited consistent changes (69% and 88%, respectively) in direction and fold-change across both mRNA and protein levels. Interestingly, previous work from our group identified significantly more transcriptomic expression changes in the DLPFC and sgPFC ^14^ than were observed for proteins in these same samples which likely due to the lower coverage of proteins afforded by mass spectrometry vs RNA-sequencing.

### Protein co-expression analysis reveals disease-specific modules and evidence for presynaptic alterations and interneuron dysfunction in PTSD

We performed protein co-expression analysis to better understand network-level protein organizational differences between PTSD and MDD. We applied WGCNA^21^ to find modules based on protein co-expression (**Fig. 2A**). PTSD-associated modules were constructed from PTSD and neurotypical control samples, and MDD-associated modules were constructed from MDD and control samples (**Extended Data Figure 3**). In total, 25 modules were identified for MDD and 28 for PTSD in DLPFC. Modules *skyblue* (correlation *r* = −0.37) and *red* (correlation *r* = 0.34) were significantly associated with PTSD diagnosis (FDR < 0.05), and modules *grey60* (correlation *r* = −0.42) and *darkred* (correlation *r* = −0.36) were significantly associated with MDD (FDR < 0.05) (**Fig. 2B**, **Extended Data Figure 3**). PTSD protein module *skyblue* is mainly composed of down-regulated proteins, including LY6H, CACNA2D1, CD59, CNTN1, and NEGR1 which has been identified as a risk gene for PTSD^12^(**Fig. 2B**). Both NEGR1 and CACNA2D1 are hub proteins in PTSD module *skyblue* and MDD module *grey60* (**Extended Data Figure 4D**).

**Figure 3.**
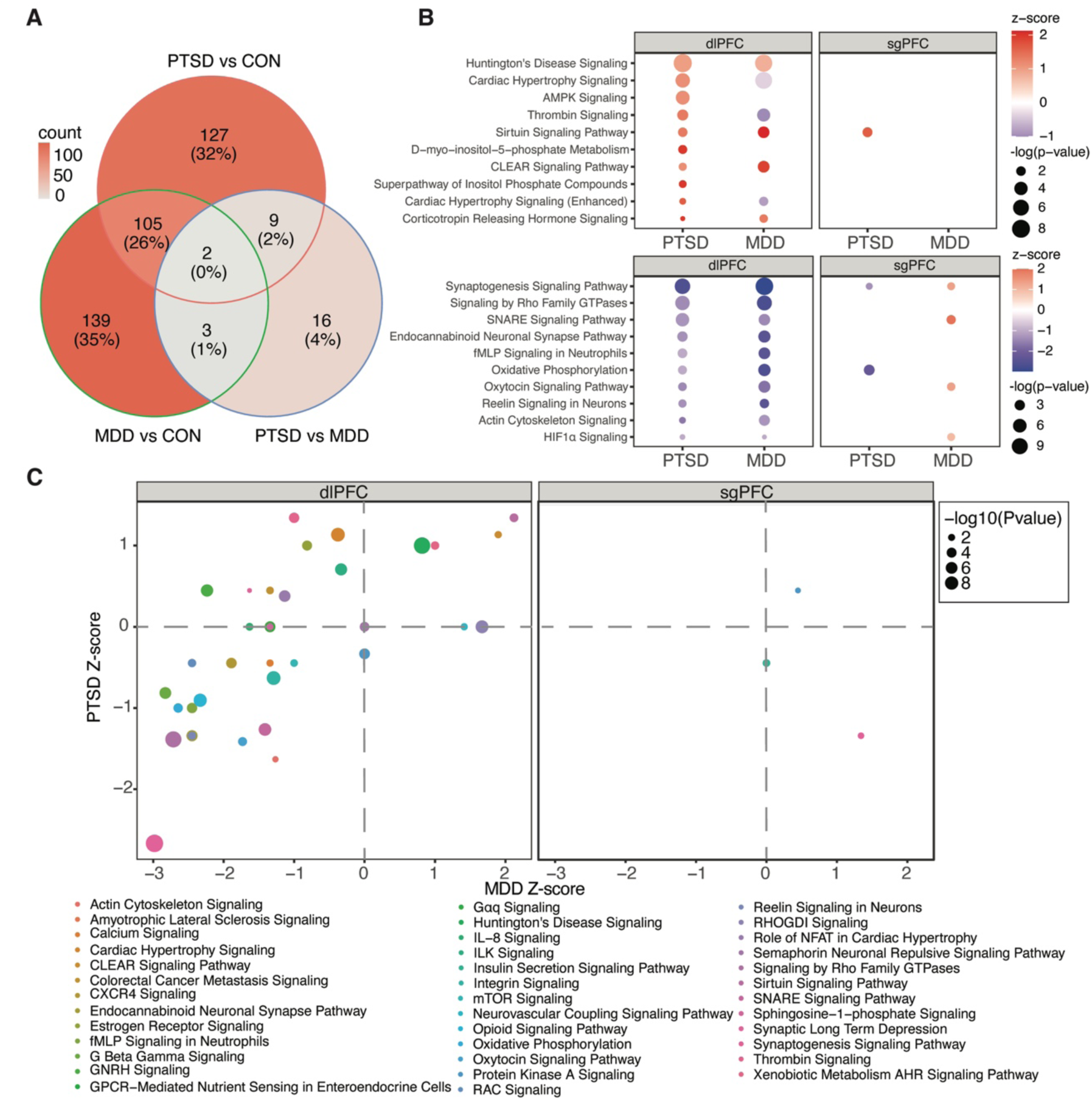
Common and divergent proteomic signatures between MDD and PTSD. (**A**) Venn diagrams show the overlap of DEPs between CON, MDD and PTSD groups in DLPFC (P<0.05). (**B**) Top significantly enriched biological pathways in DLPFC and sgPFC of PTSD and MDD datasets. (**C**) Comparison of the changing gene set enrichment (Z-scores) and directions of the biological pathways between MDD and PTSD in DLPFC and sgPFC. In DLPFC, MDD and PTSD share similar changes of pathways with an *R*^2^ = 0.57 while in sgPFC only three pathways were shared. Colors indicate gene set and circle size indicates -log10 (p-value).

**Figure 4.**
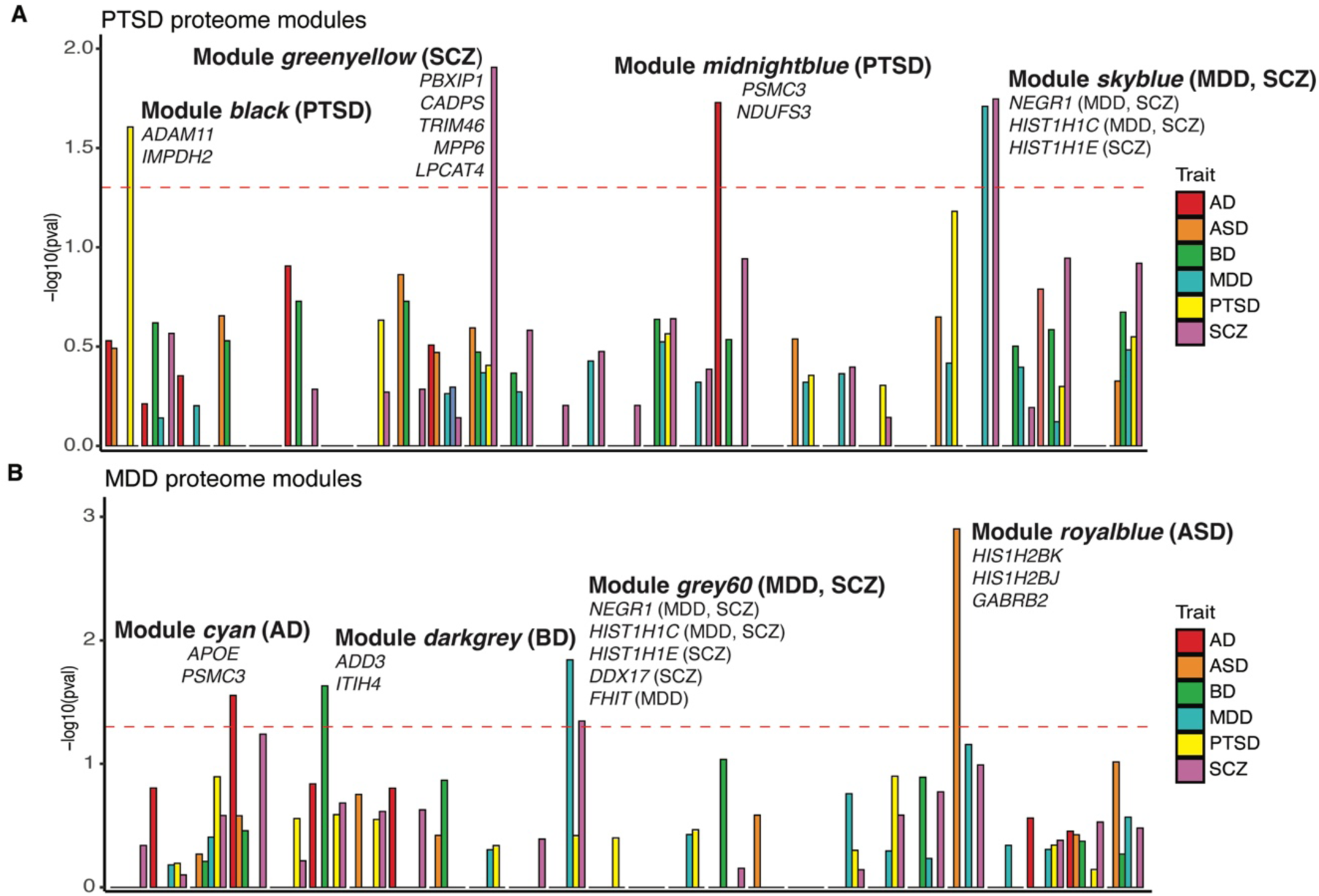
Genetic risk variant enrichment of multiple psychiatric diseases within PTSD and MDD proteomic modules. Enrichment of GWAS risk genes from TWAS analysis in the proteomic modules of PTSD (**A**) and MDD (**B**), including Alzheimer’s disease (AD, magenta bars), autism spectrum disorder (ASD, orange bars), bipolar disorder (BP, green bars), major depression disorder (MDD, blue bars), post-traumatic stress disorder (PTSD, yellow bars) and schizophrenia (SCZ, purple bars).

We next used cell type-specific enrichment analyses (CSEA^22^) to identify cell types associated with functionally distinct protein modules. CSEAs uses a cell type-specific reference profile generated with translating ribosomal affinity purification (TRAP) from transgenic BACarray reporter mice. Nine PTSD protein modules were found to be enriched for neuronal markers, including modules *skyblue* and *red* in DLPFC (**Fig. 2C**). We also observed enrichment of cell type markers for astrocytes (2 modules), oligodendrocytes (1 module) and endothelial cells (1 module) (**Fig. 2C**).

Because protein function is largely based on its location in the cell, we hypothesize that PTSD DEPs would localize to common compartments of the cell. Synaptic compartment enrichment analysis with synGO^23^ showed that modules *skyblue* and *grey60* were enriched for proteins localized to the presynaptic compartment (**Extended Data Figure 5A&B, Supp Table 2**). The PTSD-associated protein module, *red*, includes GABAergic interneuron proteins including SLC32A1 and PACSIN1, which are also members of MDD module *darkred* (**Extended Data Figure 4**). SLC32A1 encodes solute carrier family 32 member 1, a GABA/glycine vesicular transporter. Decreased protein levels of SLC32A1 is consistent with previously reported decreases in its transcript levels in PTSD frontal cortex^14^. PACSIN1 (Protein Kinase C and Casein Kinase Substrate in Neurons 1 or syndapin) plays a key role in regulating synaptic development in hippocampal interneurons ^24,25^, and mediating the modulatory effect of antipsychotic drugs in response to NMDA and glutamate^26^. Pathway analysis showed modules *red* and *darkred* were also enriched for proteins in the presynaptic compartment (adjusted P-value 1.54×10^-6^ and 5.94×10^-5^, respectively) (**Extended Data Figure 5C&D**).

**Figure 5.**
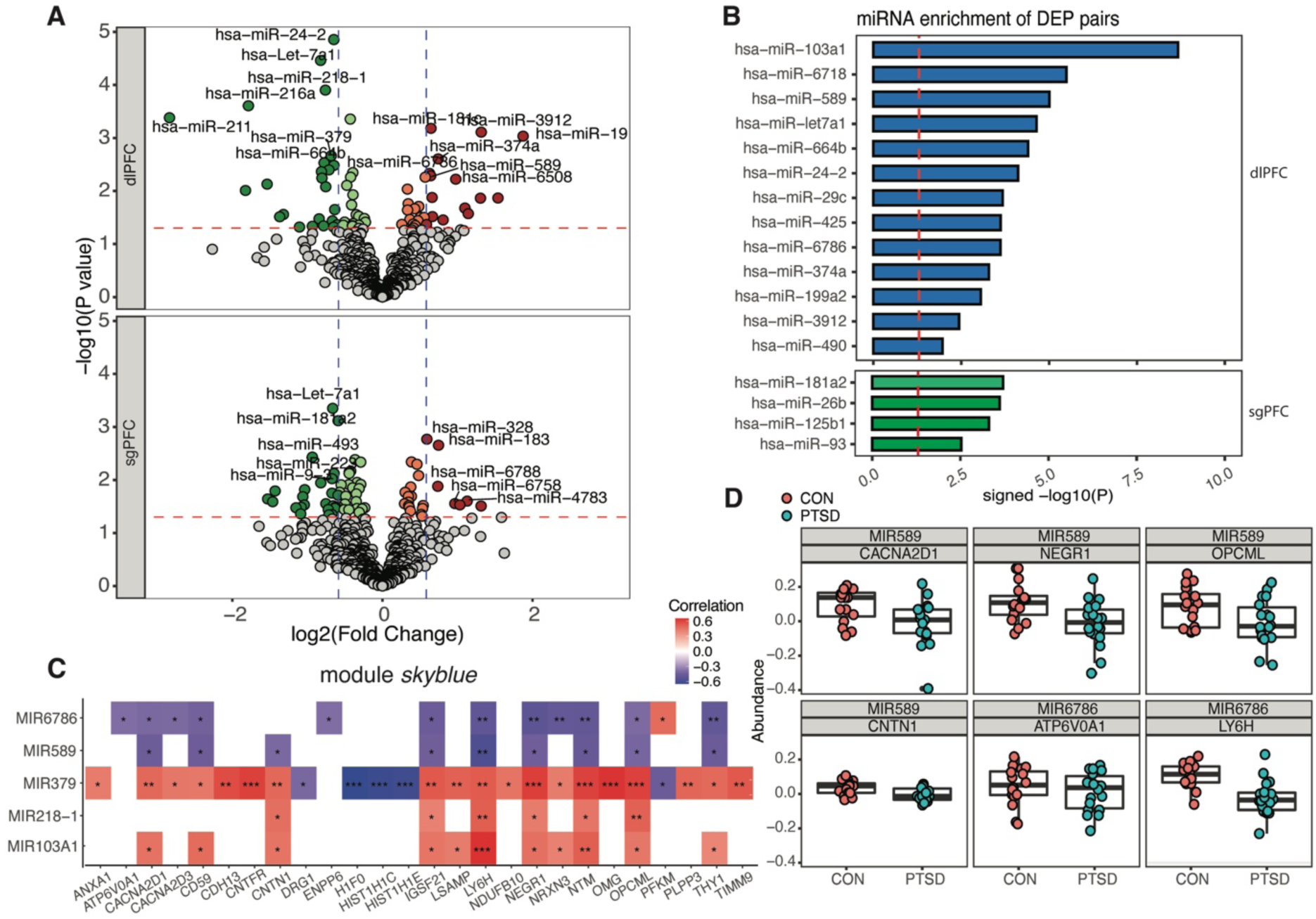
miRNA dysregulation in PTSD and its effect on the proteome. (**A**) Volcano plots showing differential expression patterns of miRNAs in response to PTSD in DLPFC and sgPFC regions. Dashed lines: log_2_(fold change) = ±0.5 and *P* value = 0.05. (**B**) miRNA enrichment analysis of DEPs to identify regulatory miRNAs and their targets. (**C**) Correlations between miRNAs with PTSD-associated protein module *skyblue*. Colors in heatmap indicate the correlation levels between miRNAs and proteins. Plotted are miRNA-protein pairs with *P* value < 0.05. (**D**) Disease-specific protein abundances (log_10_ Intensity) of putative *hsa-mir-589* targets (CACNA2D1, NEGR1, OPCML, CNTN1), and *hsa-mir-6786* targets (ATP6V0A1, LY6H) in module *skyblue*.

**Table 2.** GWASs of neurodegenerative and psychiatric diseases.

| **Disease/Disorders** | **Reference** | **Case** | **Control** | **N total** | **Loci identified** |
| --- | --- | --- | --- | --- | --- |
| Autism Spectrum Disorder (ASD) | Grove et al. 2019 | 18,381 | 27,969 | 46,350 | 12 |
| Alzheimer's Disease (AD) | Kunkle et al. 2019 | 35,274 | 59,163 | 94,437 | 25 |
| Bipolar disorder (BD) | Stahl et al. 2019 | 20,352 | 31,358 | 51,710 | 30 |
| Major Depressive Disorder (MDD) | Howard et al. 2019 | 246,363 | 561,190 | 807,553 | 102 |
| Posttraumatic Stress Disorder (PTSD) | Stein et al. 2021 | 48,221 | 217,223 | 265,444 | 18 |
| Schizophrenia (SCZ) | Pardinas et al. 2018 | 11,260 | 24,542 | 35,802 | 145 |

### Unique differential protein expression patterns between MDD and PTSD

The genetic overlap between PTSD and MDD is high (approximately 70-80%)^27^, however recent postmortem transcriptomic evidence suggests distinct molecular pathologies between the two disorders^14,16^. Parallel analysis of DEPs in MDD and PTSD postmortem brain tissue revealed moderate levels of overlap between the two diseases (**Fig. 3A**). In DLPFC, 107 DEPs (44%) overlapped between the 243 DEPs for PTSD and 249 for MDD. While far fewer protein changes were observed in sgPFC, a similar trend was observed, with 19 overlapping DEPs (33%) between the 69 DEPs for PTSD and 57 for MDD in sgPFC.

We performed gene set enrichment analyses to identify convergent and divergent biological functions associated with PTSD and MDD in each brain region. We used a less conservative significance threshold to define PTSD and MDD DEPs (nominal *P*-value < 0.05) and directionality to perform these analyses. Pathway enrichment analysis was performed using Ingenuity Pathway Analysis (IPA)^28^. Differential expression of biological pathways demonstrated shared and distinct patterns between different brain regions and disorders. In DLPFC, we noted down-regulation of pathways related to synaptic signaling (*P-*value = 4.90 × 10^-9^) and endocannabinoids (*P-*value = 8.51 × 10^-4^)), consistent with DEP enrichment in neurons (**Fig. 3B**) and previous findings that identified proteomic and transcriptomic enrichment in the synapse (**Extended Data Figure 5**) and neuronal signaling and axonal terms^14^. Corticotropin releasing hormone (CRH) signaling was up-regulated in PTSD, consistent with previous studies implicating HPA axis disruption^29^ and genetic risk^12,13^. It is interesting to note that cardiac hypertrophy signaling was up-regulated in PTSD and down-regulated in MDD (**Fig. 3B**, **Supp Table 3)**. Most converging pathway changes occurred in DLPFC (**Fig. 3C**). A full list of enriched pathways for both disorders can be found in **Supp Table 3.**

Despite the high comorbidity between MDD and PTSD, we detected many differences in the protein profiles of these two disorders (>50% of DEPS for each region). When PTSD was directly compared to MDD (not relative to a control cohort but to each other), 30 DEPs were found between the two groups including GBB4 and MACD1. Both were also identified as DEPs for PTSD vs CON and MDD vs CON comparisons, suggesting they have opposing roles in each disorder (**Supp Table 1**). In DLPFC, pathway enrichment analysis of DEPs (P<0.05) revealed dysregulation of glutamate biosynthesis (*P* = 2.52×10^-3^) as a top pathway. In sgPFC, we identified enrichment of PTSD DEPs in ketolysis (*P* = 9.81×10^-4^) and ketogenesis (*P* = 1.20×10^-3^) while the MDD-specific DEPs (*P* < 0.05) were enriched in creatine-phosphate biosynthesis (*P* = 1.34×10^-4^) and androgen signaling (*P* = 4.15×10^-4^) pathways (**Supp Table 3**). These data suggest there are differences in the proteomes of the PTSD and MDD frontal cortex that center predominantly on metabolism and neurotransmitter processing.

### PTSD proteomic modules are enriched for neuropsychiatric and neurodegenerative traits

Previous studies have reported genetic overlap between PTSD and MDD with other neuropsychiatric disorders ^14,30^. We therefore tested for enrichment of genetic risk for other neurological and neuropsychiatric disorders in PTSD protein co-expression modules (**Fig. 4A**) to identify common biological processes across brain disorders. These included Alzheimer’s disease (AD)^31^, autism spectrum disorder (ASD)^32^, bipolar disorder (BD)^33^, and schizophrenia (SCZ)^34^, along with MDD^35^ and PTSD^12^. The details of the specific GWAS summary statistics used are listed in **Table 2**. We used UTMOST (unified test for molecular signatures)^36^, a cross-tissue transcriptome-wide association algorithm based on genotype-tissue expression (GTEx)^37^, to infer disease-specific genes implicated from GWAS summary statistics of each trait. For each protein module in PTSD (**Fig. 4A**) and MDD (**Fig. 4B**), we calculated an enrichment score for the top UTMOST-inferred genes with Fisher’s exact test. PTSD module black was enriched for PTSD risk signals. Interestingly, we found PTSD module *skyblue* was enriched for both MDD (*P-*value = 0.020) and SCZ (*P-*value = 0.018) TWA genes, including NEGR1, HIST1H1C, and HIST1H1E proteins. We also identified one protein module (*midnightblue*) with highly significant enrichment (*P-*value = 0.019) for AD risk variants including PSMC3 and NDUFS3. MDD module *grey60* was enriched in MDD and SCZ risk and included many of the same proteins as PTSD module *skyblue*. We also identified MDD modules with significant enrichment for AD (module *cyan*), bipolar disorder (*darkgrey*) and autism spectrum disorder (*royalblue*) (**Fig. 4B**).

### Differential expression of miRNAs in PTSD brain

To investigate the upstream regulation of proteomic signatures resulting in PTSD, we measured the abundance of miRNAs for the same cohort including 53 samples from DLPFC and 55 samples from sgPFC (**Table 1**). We generated RNA sequencing libraries enriched for small non-coding RNAs (smRNA-seq) derived from total RNAs for analysis (**Extended Data Figure 6A**). We found notable differences in both coverage and total expression levels of miRNAs between bulk RNA-seq of total RNAs and small RNA-seq of non-coding RNAs. 1,365 miRNAs passed quality control, mapping, and filtering criteria for both brain regions. smRNA-seq provided higher coverage and deeper sequencing depth of shorter transcripts, especially miRNAs, than those from total RNAs, irrespective of region and diagnosis (see **Extended Data Figure 6B**). Supervised hierarchical clustering analysis of these miRNA samples revealed stronger local clustering patterns for primary diagnoses (PrimaryDx) and brain region than other demographic features (e.g., sex, age at death, ancestry) and technical sequencing variables (RNA integrity number (RIN), postmortem interval (PMI)) (**Extended Data Figure 6C**), consistent with Principal Component Analysis (PCA) results revealing high correlations with top PCs with brain region (**Extended Data Figure 6D**). Next, we performed differential expression analysis of miRNA datasets to identify changes in PTSD cases vs. controls and in DLPFC, we identified expression changes for six miRNAs. In the PTSD sgPFC, we identified two differentially expressed miRNAs and 19 in MDD (2 overlapping with PTSD) (**Fig. 5A**, **Supp Table 4)**. Analysis of differential miRNA expression revealed brain region-specific patterns (**Extended Data Figure 7A**), with relatively low overlap between the diagnostic cohorts (19% in DLPFC and 34% in sgPFC) using a nominal cutoff of *P* < 0.05 (**Extended Data Figure 7B**). Six PTSD miRNAs including *hsa-mir-24-2*, *hsa-let-7a1*, *hsa-mir-218-1*, *hsa-mir-216a*, *hsa-mir-211*, *hsa-mir-425* passed FDR threshold and three miRNAs did for MDD, *hsa-mir-589*, *hsa-mir-6508*, *hsa-mir-655*.

### Disruption of miRNA co-expression modules in PTSD

To investigate the organization of miRNA expression, we performed network analysis based on the co-expression profiles of miRNAs using WGCNA^21^ (**Extended Data Figure 7C**). We identified 6 miRNA co-expression modules with significant demographic trait relationships (**Extended Data Figure 7C**). One miRNA co-expression module, *turquoise*, had significant association with PTSD (**Extended Data Figure 7D**), indicating the enrichment of PTSD-associated signals in this module. We identified 18 up-(red) and 7 down-regulated (green) miRNAs within module *turquois*e with a nominal *P*-value < 0.05 (**Extended Data Figure 7E**). Among the most significantly regulated miRNAs was *hsa-mir-589* (up regulated 1.56 fold in dlPFC), which has previously been implicated in MDD pathophysiology^38^.

### PTSD proteomic regulation by miRNAs

Because miRNAs are known to regulate protein translation (normally inhibiting it), we hypothesized that protein-mRNA pairs with fold changes in opposite directions in PTSD (**Fig. 1D**) might be regulated by disease-associated miRNAs. With further investigation of these proteins we found that all four DLPFC proteins (DPP6, CD59, RHOB, and PTPRE) and two out of three sgPFC proteins (CD59 and RHOB) were associated with both expression and target 3’-UTR sequences of PTSD regulated miRNAs (**Supp Table 5**).

To better understand the relationship between protein expression and miRNA function we integrated the two datasets. First, we performed *Pearson’s* correlation test between the normalized expression levels of the protein (in log_10_LFQ intensity) and the miRNA (in log_2_FPKM). An example is provided in **Extended Data Figure 8A** for proteins SLC32A1 and LY6H with miRNA *hsa-mir-589*. Second, for each miRNA, we calculated an enrichment score for DEPs among the proteins associated with that miRNA using Fisher’s exact test. Using this approach, we identified miRNA dysregulations that were PTSD-or MDD-specific or associated with both disorders (**Extended Data Figure 8B,C**). In DLPFC, 21 miRNAs were enriched for PTSD DEPs (**Fig. 5B**), MDD DEPs (**Extended Data Figure 8B**) or both. And in the sgPFC four miRNAs were enriched for PTSD DEPs (**Extended Data Figure 8C**). The miRNA with the most significant enrichment for PTSD-specific DEPs was *hsa-mir-103a1* which in blood plasma samples was down-regulated in patients with childhood traumatization^39^. The miRNAs that were most significantly enriched for both MDD and PTSD DEPs included *hsa-mir-6786* (P=3.22×10^-6^ (PTSD) and 1.38×10^-11^ (MDD)) and *hsa-mir-589* (P=9.85×10^-6^ (PTSD) and 3.95×10^-9^ (MDD)). While little is known about the function of *hsa-mir-6786*, *hsa-mir-589* has been previously implicated in depression^40^, and it was previously identified in our PTSD miRNA co-expression *turquoise* module (**Extended Data Figure 7E**). In addition, one of its predicted targets from TargetScan is the GABA vesicular transporter protein SLC32A1, which is significantly downregulated in PTSD DLPFC (**Fig. 2B**).

We identified nine miRNAs exhibiting significant enrichment for associated DEPs (nominally significance of *P* < 0.05) in sgPFC. There were far fewer miRNAs with significant enrichment of DEPs in the sgPFC. *hsa-mir-181a2* was the most significantly enriched miRNA for PTSD DEPs (P=1.94×10^-4^) and *hsa-mir-24-2* (P=2.81×10^-5^) and *hsa-mir-9-3* (P=3.41 × 10^-5^) had the most significant enrichment MDD-specific DEPs. In addition, *hsa-mir-125b1* was enriched for both MDD and PTSD DEPs (P=3.44×10^-3^ and 4.90×10^-4^, respectively).

### miRNA-regulated protein co-expression modules

We next used Fisher’s exact test to determine the enrichment of proteins within particular modules among proteins that were co-expressed with a given miRNA to further explore the relationship between differentially expressed miRNAs and proteins under their regulation. A significant enrichment score (*P*-value < 0.05) indicated the overrepresentation of module members with proteins connected with a particular miRNA. In other words, proteins correlated with the miRNA have a significantly higher probability of coming from the specified protein module, indicating the likelihood of co-regulation of the local protein network by the miRNA. The full miRNA-module enrichment results are annotated in **Extended Data Figure 9A**.

Five differentially expressed miRNAs were significantly correlated with protein levels in the PTSD-associated module *skyblue* (**Fig. 5C**), indicating a potential regulatory relationship between the miRNAs and the protein members. Among them, *hsa-mir-6786* and *hsa-mir-589* were negatively correlated (*P* < 0.05) with the majority of proteins in module *skyblue* (21 out of 22 pairs). We compared our miRNA-protein pairs with those predicted by TargetScan^41,42^ and found that the DEPs LY6H and ATP6V0A1 are targets of *hsa-mir-6786*. TargetScan also identified six targets present in module *skyblue* that were negatively associated with *hsa-mir-589* expression, including CACNA2D1, CNTN1, THY1, OPCML, CD59, and NEGR1 (**Fig. 5C**, **Supp Table 6**). Because miRNAs generally act to reduce translation of proteins, we would predict negative associations between miRNAs and their regulated protein targets. Both CACNA2D1 and CNTN1 were also confirmed in an independent miRNA database MirBase^43–48^ as targets of *hsa-mir-589*. Shared and unique miRNAs were enriched for MDD modules as well. In MDD module *grey60*, six miRNAs were significantly enriched (P<0.05), and three were negatively correlated with expression of eight out of nine pairs of the proteins in module *grey60*, including *hsa-mir-6786*, *hsa-mir-589*, and *hsa-mir-4745* (**Extended Data Figure 9B**). Both *hsa-mir-6786* and *hsa-mir-589* were shared with PTSD. Similar to PTSD module *skyblue*, MDD module *grey60* included predicted targets of *hsa-mir-6786*, including ATP6V0A1, GPC1, ICAM5, and LY6H, and those of *hsa-mir-589*, including CACNA2D1, NEGR1, OPCML, CD59, LSAMP, and SH3GLB2. The regulatory pairs of *hsa-mir-589*-CACNA2D1 and *hsa-mir-6786*-ICAM5 were also identified by MirBase. MDD module *grey60* also included the predicted targets of *hsa-mir-4745*, including CNTFR, CD59, NEGR1, and SH3GLB2. In the MDD module *darkred*, only *hsa-mir-4745* was found to be enriched and negatively correlated with 8 of its associated proteins (**Supp Table 6**). This module contained predicted targets of *hsa-mir-4745*, including GPRIN1, PACSIN1 and MAPT (**Extended Data Figure 4**). Among these, the regulation of PACSIN1 by *hsa-mir-4745* was confirmed in MirBase. Taken together, these analyses identify specific miRNA-protein pairs (*hsa-mir-589*-CACNA2D1/CNTN1 and *hsa-mir-6786*-LY6H/ATP6V0A1) that likely play significant roles in PTSD pathophysiology (**Fig. 5D**).

## Discussion

In this study, we present a comprehensive evaluation of the human prefrontal cortex neuroproteome in neurotypical control, MDD and PTSD donors. Our investigation centered on two frontal cortical regions implicated in stress disorders: the DLPFC and sgPFC. To our knowledge this is the largest postmortem case/control proteomics cohort for PTSD to date. By integrating protein, mRNA and miRNA profiles from the same samples, we more deeply characterize mechanisms that control PTSD and MDD-related alterations in protein levels associated with the pathophysiology of these disorders. In addition, we integrated genome wide-association studies of other neuropsychiatric and neurodegenerative disorders to identify common proteomic signatures with PTSD.

One model of PTSD neurobiology suggests top-down impairment of the fear circuitry of the brain^49^. Decreased prefrontal cortical activity has been observed in PTSD patients^50–54^ and this likely results in the hyper-activation of subcortical regions (e.g. amygdala) resulting in exaggerated responses to fearful stimuli, a hallmark of PTSD. PTSD-associated protein module, *red*, includes the down regulated interneuron protein SLC32A1, a synaptic GABA transporter. Downregulation of *SLC32A1* has been reported previously in transcriptomic studies of the prefrontal cortex of the PTSD brain^14^. Further, *SLC32A1* transcript is a putative target of *hsa-mir-589* (**Supp Table 4**). We previously found that *SLC32A1* transcript was closely connected with expression of other interneuron molecular key drivers in PTSD postmortem brain tissues and was downregulated in the DLPFC. *ELFN1* was identified as a significant TWAS hit and a significant differential methylation site in subcortical regions of PTSD brain^55^, indicating genetic regulation of GABAergic transcripts in PTSD brain. ELFN1 organizes and transports synaptic components of interneurons (including mGLUR7) to the post synaptic density of GABAergic neurons^56^. The MDD-associated neuronal module, *darkred*, contained 50 DEPs, including PACSIN1 and MAPT. These two proteins together regulate microtubule cytoskeleton reorganization in neuronal axons and synapses^57,58^. Both the PTSD modules *skyblue* and *red*, and MDD modules *grey60* and *darkred*, are also significantly enriched for proteins specific to the presynaptic compartment (**Extended Data Figure 5**) including LY6H which regulates neurotransmitter trafficking in neurons^59^. Further, *hsa-mir-589* expression is negatively correlated with several proteins in these modules including CACNA2D1, CACNA2D3, NEGR and OPCML and as previously noted, regulates expression of the GABA transporter SLC32A1 which is significantly down-regulated in PTSD. We hypothesize that *hsa-mir-589* acts through its effector proteins, which are involved in neuronal plasticity and interneuron function, to disrupt synaptic transmission in the prefrontal cortex to other fear centers of the brain. Taken together these findings point to impaired GABAergic function in the cortex of PTSD patients and represent one possible molecular mechanism by which prefrontal activity inhibition manifests in PTSD.

Significant progress has been made to identify cross-disorder overlap in the genomic signals among psychiatric disorders. Recently, Gandal, et al.^30^ identified significant correlations in the transcriptomes of prefrontal cortical regions among schizophrenia (SCZ), autism spectrum disorder (ASD), and bipolar disorder (BD) and some overlap with MDD and alcohol use disorder. In our previous work^14^, we compared the DLPFC transcriptome of PTSD to these same disorders and after uniform processing of our data, identified significant transcriptomic correlation between PTSD and SCZ, ASD and bipolar disorder. However, our findings did not identify significant correlation between PTSD and MDD. In this study, we likewise found only moderate overlap in DEPs and regulated miRNAs between the two disorders but there was some convergence in protein co-expression patterns. We identified brain region- and disease-specific protein co-expression modules for PTSD and MDD. Both the PTSD module *skyblue* and MDD module *grey60* are also enriched for SCZ and MDD GWAS signals (**Fig. 4**). The PTSD-associated neuronal module, *skyblue*, contains 30 proteins, including downregulated DEPs, CACNA2D1, OPCML, and NEGR1, which are shared by the MDD-associated neuronal module *grey60*. OPCML is an opioid receptor that has been previously reported as a PTSD risk gene^12^. OPCML has also been shown to associate with susceptibility for both SCZ and MDD^60,61^. NEGR1 regulates neural growth^62^ and has been reported as a genetic risk gene for MDD^63–66^ and other comorbid disorders such as anxiety^67^ and eating disorders^68^. Both OPCML and NEGR1 similarly have functions in neural cell adhesion in the cortex of human SCZ patients^69^. CACNA2D1 encodes the *α*_2_*δ* subunit of the voltage-gated calcium channel and is one of the most replicated risk genes for SCZ^70^. These findings point to common disruption of cell adhesion and neural connectivity processes among PTSD, MDD and SCZ.

PTSD and Alzheimer’s Disease are highly comorbid^71,72^ but it is still unclear if and how PTSD is a risk factor for developing AD^73^. Nevertheless, two major findings from our study offer insight into the molecular intersections between PTSD and AD pathology. First, we identified a PTSD protein co-expression module *midnightblue* enriched for AD risk genes (PSMC3, NDUFS3). Second, we found significant PTSD-expression changes in *hsa-mir-24* and *hsa-mir-125b1*, both of which are altered in AD brain. *In silico* analysis of postmortem brain found *hsa-mir-24* was up-regulated in AD^74^ and down-regulated in AD blood serum^75^. *hsa-mir-24* also regulates microglia polarization after traumatic brain damage in rodent models^76^. Interestingly, the *hsa-mir-125B1* gene is differentially hypermethylated in AD versus control temporal cortex tissue but could not be linked to differential expression of *hsa-mir-125b1* transcript ^77^. Previous longitudinal studies of depression phenotypes in aging cohorts have also identified global miRNA expression changes^78^ (including miRNAs *hsa-mir-484-5p*, *hsa-mir-26b-5p*, *hsa-mir-30d-5p*, and *hsa-mir-1973p*) that are associated with late-life depression and a higher probability of developing Alzheimer’s dementia. While the current study could not confirm changes in these specific miRNAs (likely due to technical variance between Nano-string and smRNA-sequencing and the mean age of each cohort (mean age 86.9 ± 6.6 years vs 46.2 ± 11.9 years)), our findings identify novel candidates for key molecular links between PTSD and AD disease etiology.

In conclusion, we present the largest human postmortem proteomics study for PTSD and MDD to date. This work fills a critical gap bridging transcriptomic and proteomic genomic signatures in postmortem molecular studies. We identified changes in key PTSD proteins and pathways including interneuron and CRH signaling including the GABA transporter SLC32A1. Surprisingly, we found only moderate overlap in the proteomic signatures between PTSD and MDD but identified co-expression patterns common to both disorders. Cross disorder analysis of our proteomics dataset with the largest genome-wide studies available also identified AD, SCZ, and MDD risk variant enrichment in PTSD co-expression networks, suggesting pathophysiological convergence and common risk for these disorders. Further, we were able to confirm many miRNA-DEP interactions that were predicted by TargetScan and miRbase including CACNA2D1, CNTN1, THY1, OPCML, CD59, NEGR1, and SLC32A1 as targets of the miRNA *hsa-mir-589*. These results confirm GABAergic impairment and synaptic dysfunction identified in previous transcriptomic and epigenetic studies of PTSD and highlight the key proteins involved, along with their paired regulatory miRNAs. In addition, this large proteomics dataset serves as a rich resource for functional genomics and translational architecture in different human cortical brain regions. Together, these efforts will lead to progress in development of novel therapeutics and treatments for PTSD.

## Online Methods

### Human Postmortem Cohort

This study was conducted using frozen postmortem brain specimens from the University of Pittsburgh Medical Center. Individuals were a mix of European, Asian, and African American descent. The cohorts were matched for sex, age, PMI, and pH. Summary of sociodemographic and clinical details are listed in **Table 1** and includes the presence of comorbid disorders, tobacco use, manner of death, and presence of drug and/or alcohol abuse. A total of 66 individuals (22 PTSD: 11 males, 11 females; 22 MDD: 11 male, 11 female; and 22 healthy controls: 11 males, 11 females). Fresh frozen tissue samples (25mg) of PFC from the sgPFC (BA 25) and the DLPFC (BA 9/46) were collected for each donor.

Psychiatric history and demographic information were obtained via an extensive psychological autopsy including: medical record collection and review; structured diagnostic interviews (SCID-5-RV and SCID-5-PD) with the next-of-kin or other knowledgeable informant; review of neuropathology and toxicology reports; and a postmortem diagnostic conference staffed by adult, child and geriatric psychiatrists, clinical psychologists and other senior psychiatric clinicians, that generates DSM-IV/5 and ICD-10-CM diagnoses (or their absence) for all subjects as previously described. Of the 22 individuals with PTSD, 50% (9 MDD and 2 depressive disorder NOS) were also comorbid for depression.

### smRNA-seq library preparation

RNA was isolated from 20mg of frozen postmortem brain tissue using a RNeasy Mini Kit with genomic DNA elimination, as described the manufacturer (Qiagen). The RIN and concentration were assessed using a Bioanalyzer (Agilent). smRNA libraries were constructed using a QiaSeq miRNA library kit (Qiagen) from 1ug of RNA. Samples were barcoded and sequenced on a HiSeq2500 (Illumina) at a read depth of 20M.

### Tissue Collection and preparation for LC-MS/MS

Tissue was lysed in RIPA buffer with protease and phosphatase inhibitors (100x halt inhibitor cocktail, Thermofisher) using a probe sonicator. Cellular debris was pelleted by centrifugation and supernatant containing soluble proteins was collected. Soluble protein (20ug in 10 ul) was aliquoted from the supernatant, additional water was added to a final volume of 100uL. Samples was then added to 200 uL of ice-cold acetone and protein was precipitated overnight. The pellet was air dried and resuspended in 20uL 8M urea, containing 400mM ammonium bicarbonate (pH 8) and reduced in dithiothreitol (DTT; 2µL, 45 mM) at 37°C for 30 min. Samples were alkylated with iodoacetamide (IAM; 2µL, 100 mM) at room temperature (in the dark) for 30 min. Additional water was added and samples were enzymatically digested with sequencing-grade trypsin (1:40 trypsin:protein; Promega, Madison, WI, USA) at 37°C for 16 hours. The final volume was 80 uL. Digested proteins were acidified in 0.1% formic acid and desalted via column purification (C18 spin columns; The Nest Group, Inc; Southborough, MA, USA) and dried using a SpeedVac. Samples were stored in −80°C until mass spectrometry analysis.

### Data Independent Analysis (DIA) Mass Spectrometry

Purified samples were resuspended in 0.2% trifluoroacetic acid (TFA)/2% acetonitrile (ACN) in water. DIA LC-MS/MS was carried out with a nano-ACQUITY UPLC system (Waters Corporation, Milford, MA, USA) connected to an Orbitrap Fusion Tribrid (ThermoFisher Scientific, San Jose, CA, USA) mass spectrometer. Samples were injected and loaded into a trapping column (nanoACQUITY UPLC Symmetry C18 Trap column, 180 µM x 20 mm) at 5µL/min. Peptides were subsequently separated using a C18 column (nanoACQUITY column Peptide BEH C18, 75 µm × 250 mm). Mobile phases consisted of Mobile Phase A (0.1% formic acid in water) or Mobile Phase B (0.1% formic acid in ACN). Peptides were eluted with 6-35% gradient Mobile Phase B for 90 min, 85% Mobile Phase B for 15 min at 300nL/min, 37°C. All sample injections were interspersed by column regeneration and three blank injections. Data were acquired under Data Independent Acquisition (DIA) mode with an isolation window of 25m/z. Full scan was in the 400-1000m/z range, “Use Quadrupole Isolation” enabled at an Orbitrap resolution of 120,000 at 200m/z and automatic gain control target value 4 x 10^5^. MS^2^ fragment ions were generated in C-trap with higher-energy collision dissociation at 28% and Orbitrap resolution of 60,000.

DIA spectra were searched against a *homo sapiens* brain proteome fractionated spectral library generated from DDA LC MS/MS spectra (collected from the same Orbitrap Fusion mass spectrometer with HDC fragmentation) using Scaffold DIA software v. 1.2.1 (Proteome Software, Portland, OR, USA). Within Scaffold DIA, raw files were first converted to the mzML format using ProteoWizard v. 3.0.11748. The samples were then aligned by retention time and individually searched with a mass tolerance of 10 ppm and a fragment mass tolerance of 10 ppm. The data acquisition type was set to “Overlapping margins of 2Da”, and the maximum missed cleavages was set to 2. Fixed modifications included carbamidomethylation of cysteine residues (+57.02). Dynamic modifications included phosphorylation of serine, threonine, and tyrosine (+79.96), deamination of asparagine and glutamine (+0.98), oxidation of methionine and proline (+15.99), and acetylation of lysine (+42.01). Peptides with charge states between 2 and 4 and 6-30 amino acids in length were considered for quantitation, and the resulting peptides were filtered by Percolator (v. 3.01) at a threshold FDR of 0.01. Peptide quantification was performed by EncyclopeDIA (v. 0.9.2), and six of the highest quality fragment ions were selected for quantitation. Proteins containing redundant peptides were grouped to satisfy the principles of parsimony, and proteins were filtered at a threshold of two peptides per protein and an FDR of 1%.

### Differentially expressed proteins and RRHO

Differential expression analysis for proteins was performed with R package *limma*. Any proteins with 0 peptides were dropped. For each brain region, an empirical Bayes linear regression model was used to fit protein expression with PrimaryDx and covariates, including the age of death, race, and sex. P values were adjusted to control FDR.

A rank-rank hypergeometric overlap (RRHO) plot was made with R package *RRHO*. For each pair of conditions, log_10_ fold changes of corresponding proteins in different conditions were compared after ranking.

### Expression calling from smRNA-seq data

smRNA-seq data in FASTQ files were mapped to the human reference genome and annotation GTF file (GRCh38, release 104) downloaded from ENSEMBL with software *STAR* (v2.5.3a) and counted with *featureCounts* (v1.5.3). In *STAR*, parameter settings included *outFilterMultimapScoreRange* set to 0, *outFilterMatchNmin* set to 16, *outFilterMatchNminOverLread* and *outFilterScoreMinOverLread* set to 0.3. In *featureCounts*, counting was conducted on fragments with exon annotations on the transcript level, stranded, and multiple assignments allowed. In total, 1,365 miRNAs were found across all samples.

### Differentially expressed miRNAs

miRNA expression counts were extracted from smRNA-seq data. Differential expression analysis for miRNAs was performed with R package *DESeq2* in a region-specific manner. In each region, miRNAs with an average count less than 0.5 across all samples were filtered out. Then expression was modeled with PrimaryDx and covariates, including the age of death, RIN (RNA integrity number), and sex.

### Pathway enrichment analysis

Pathway enrichment analysis of DEPs was performed in IPA (QIAGEN Inc., https://www.qiagenbioinformatics.com/products/ingenuity-pathway-analysis). A cutoff of P-value < 0.05 was used to define significantly changed pathways. Neuronal cell compartment pathway enrichment analysis of protein modules was conducted with SynGO: https://www.syngoportal.org/index.html.

### Protein co-expression modules and network analysis

Protein co-expression modules were constructed with the R package *WGCNA*. Control and MDD samples were used to construct MDD modules; control and PTSD samples were used to construct PTSD modules. The module-trait correlation was calculated between module eigengenes and sample traits, including PrimaryDx, age of death, PMI (postmortem interval), race, and sex. Soft-threshold powers (6 for DLPFC conditions and 4 for sgPFC conditions) were used to achieve approximate scale-free topology in each condition. Protein modules were built with the *blockwiseModules* function in *WGCNA*. Modules were labeled with random colors. P values were FDR-corrected to adjust for multiple comparisons. *TOMtype* was set to “signed” and *mergeCutHeight* was set to 0.1. Minimum of module sizes was set to 20.

Protein-protein interaction networks were inferred from mutual information with protein expression using algorithm ARACNE^79^ implemented by R package *bnlearn*. The network organization of modules was visualized using the R package *igraph*. Here each node represents a protein, and the lines between them show significant protein-protein co-expression. Finally, undirected key driver analysis was performed in the local networks of protein modules with R package *KDA* (https://labs.icahn.mssm.edu/binzhanglab/resources/) to identify key driver proteins of the corresponding module.

### Cell type-specific enrichment analysis (CSEA)

R package *pSI* (http://genetics.wustl.edu/jdlab/psi_package/) was used to perform cell type-specific enrichment analysis for each module (either MDD or PTSD). Cell type-specific gene expression profiles included neurons, astrocytes, microglia, oligodendrocytes, and endothelial cells, based on human data from Gene Expression Omnibus (GEO) with accession ID GSE73721. Gene expression was log-transformed, and mean values were calculated for each cell type. Cell type-specific enrichment for the WGCNA module was conducted with function *specificity.index* in the *pSI* package. Fisher’s exact test was used to test the significance with a pSI threshold of 0.05. The obtained P values were FDR-corrected.

### Module preservation between MDD and PTSD

For each brain region and a given pair of MDD and PTSD modules, their module consistency was calculated using Fisher’s exact test to look for the overrepresentation of proteins of a paired module in the other. P values were FDR-corrected, and module pairs were significantly preserved if FDR < 0.05. The enrichment score is represented by the odds ratio of protein overlap over the expected numbers.

### Comparison between transcriptomics and proteomics

The consistency of transcriptomic and proteomic changes was compared. For each brain region and disorder, genes with a P-value < 0.05 for disease association on both RNA and protein levels were selected. Log_2_ fold changes (log_2_(FC)) were compared for each gene. Transcriptomic log_2_(FC) were obtained from the previous transcriptomic study of MDD and PTSD from the same brain region^14^.

### Protein-miRNA pairs

Correlations of protein-miRNA pairs were calculated based on protein and miRNA expression levels from the same individuals. Samples were selected with both proteomics and miRNA measurement. For control-PTSD, there were 27 matched samples in DLPFC and 31 in sgPFC; for control-MDD, there were 14 in DLPFC and 33 in sgPFC. miRNAs with average raw expression > 0.5 were kept and transformed to FPKM. A correlation test was performed for each protein and miRNA pair with protein log_10_(intensity) and miRNA log_2_(FPKM). A significant protein-miRNA connection is defined by P-value < 0.05.

### miRNA enrichment of DEPs and protein modules

miRNAs’ enrichment of DEPs and protein modules was estimated based on protein-miRNA pairs described above. For each miRNA, the enrichment score was the odds ratio of overrepresentation of DEPs (defined by protein-disease association P-value < 0.05) among all the proteins connected with the miRNA. A Fisher’s exact test was applied to the enrichment to get a P-value. Significant DEP enrichment was defined by a P-value < 0.05.

miRNA-module enrichment was done similarly. For a given miRNA and a protein module, the enrichment score was the odds ratio of overrepresentation of the module membership among all the proteins connected with the miRNA. P values were obtained from Fisher’s exact test. Significant enrichment of protein modules was defined as P-value < 0.05.

### Protein module enrichment of multi-trait TWAS results

GWAS summary statistics were downloaded from studies listed in **Table 2**. TWAS predictions for each trait were made with UTMOST by joint analysis of individual predictions from all brain tissues listed in GTEx release v6p. UTMOST performs gene expression imputation across tissues and gene-level association tests to identify trait-associated genes from GWAS summary statistics. The complete list of 44 tissues is on the GTEx website (https://gtexportal.org/home/). The top 100 predicted genes were used for each trait to balance different psychiatric disorders. A Fisher exact test was performed for each protein module to test for the overrepresentation of top UTMOST predicted genes in the module among all genes. Significant enrichment was marked by a P-value < 0.05.

### Code Availability

All code used in this study is freely available online and can be found at https://github.com/mjgirgenti/.

### Data Availability

Mass Spectrometry and RNA-seq summary statistics and raw data generated and/or analyzed during the current study will be made available on Gene Expression Omnibus.

## Data Availability

All data produced in the present study are available upon reasonable request to the authors

## Extended Data Figures

**Extended Data Fig 1.**
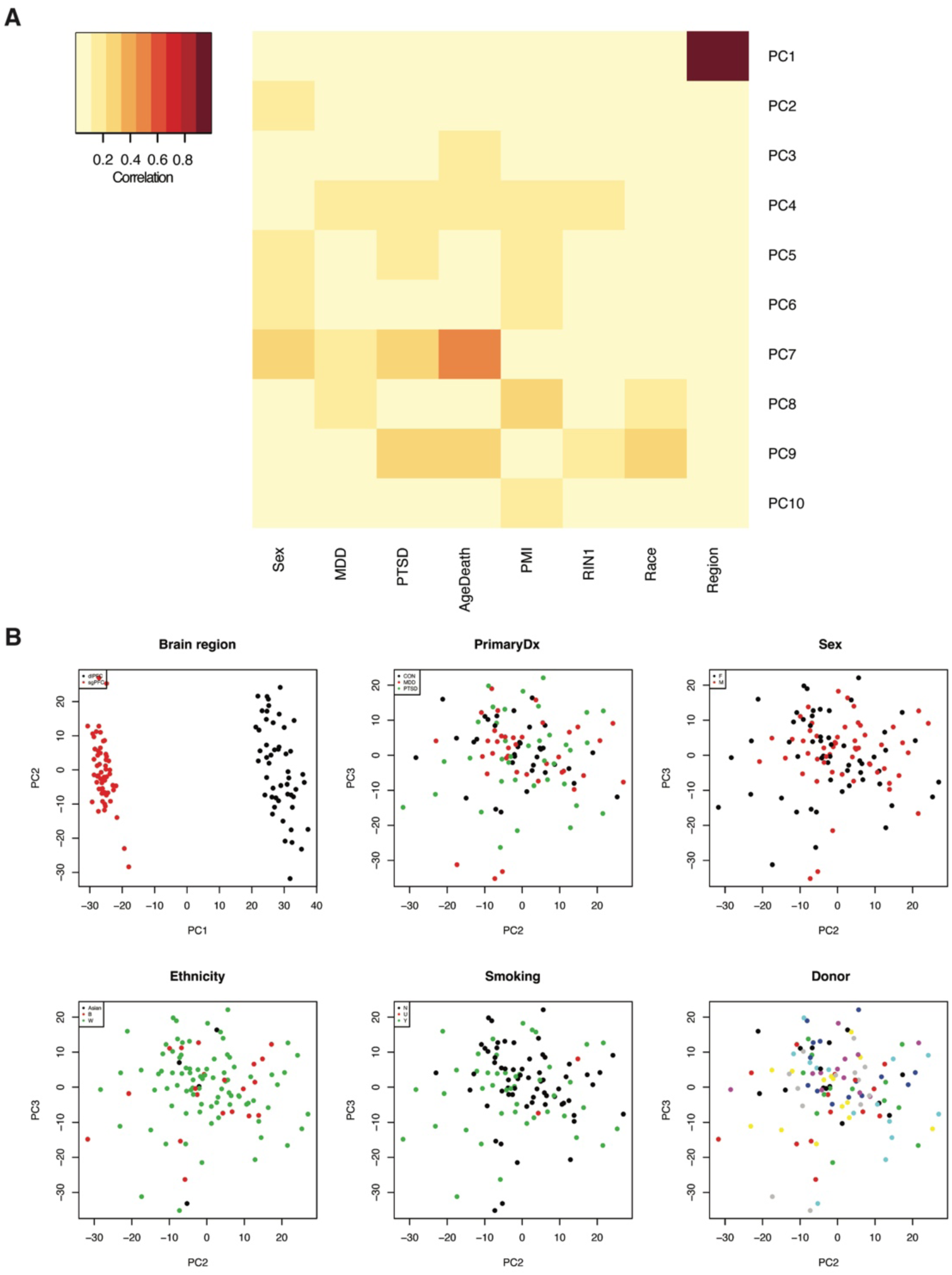
Quality control and PCA of proteomics samples. (**A**) Correlations between top PCs and traits. (**B**) Effects of selected traits in top PC space.

**Extended Data Fig 2.**
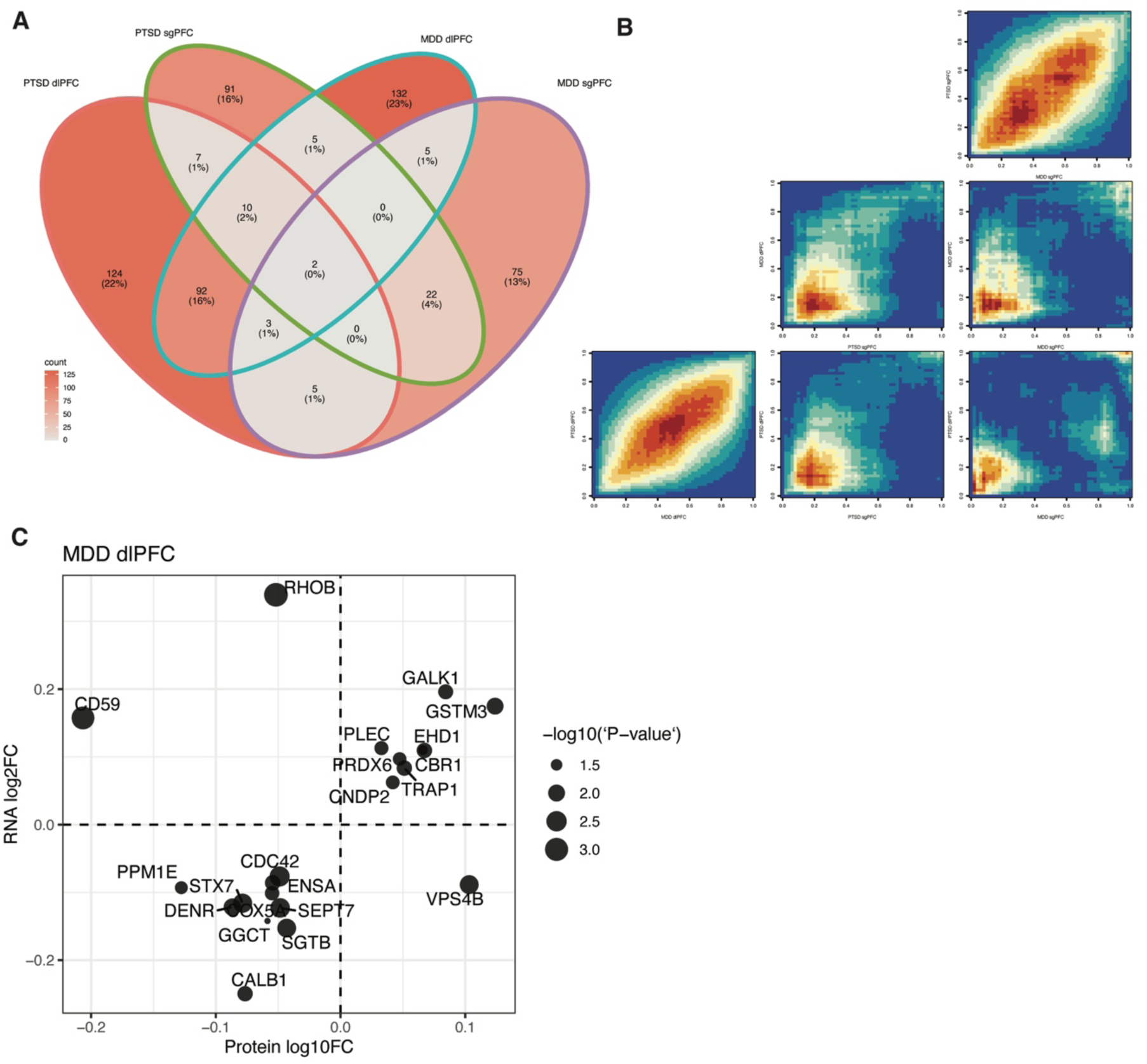
Protein differential expression analysis. (**A**) Venn diagram showing overlap of DEPs (nominal *P* < 0.05) between regions and diagnostic cohorts. (**B**) RRHO of protein differential expression. (**C**) Convergence between transcript and protein differential expression in DLPFC of MDD brains (Adjusted *R*^2^ = 0.096, robust regression *P* = 7.59 × 10^-7^).

**Extended Data Fig 3.**
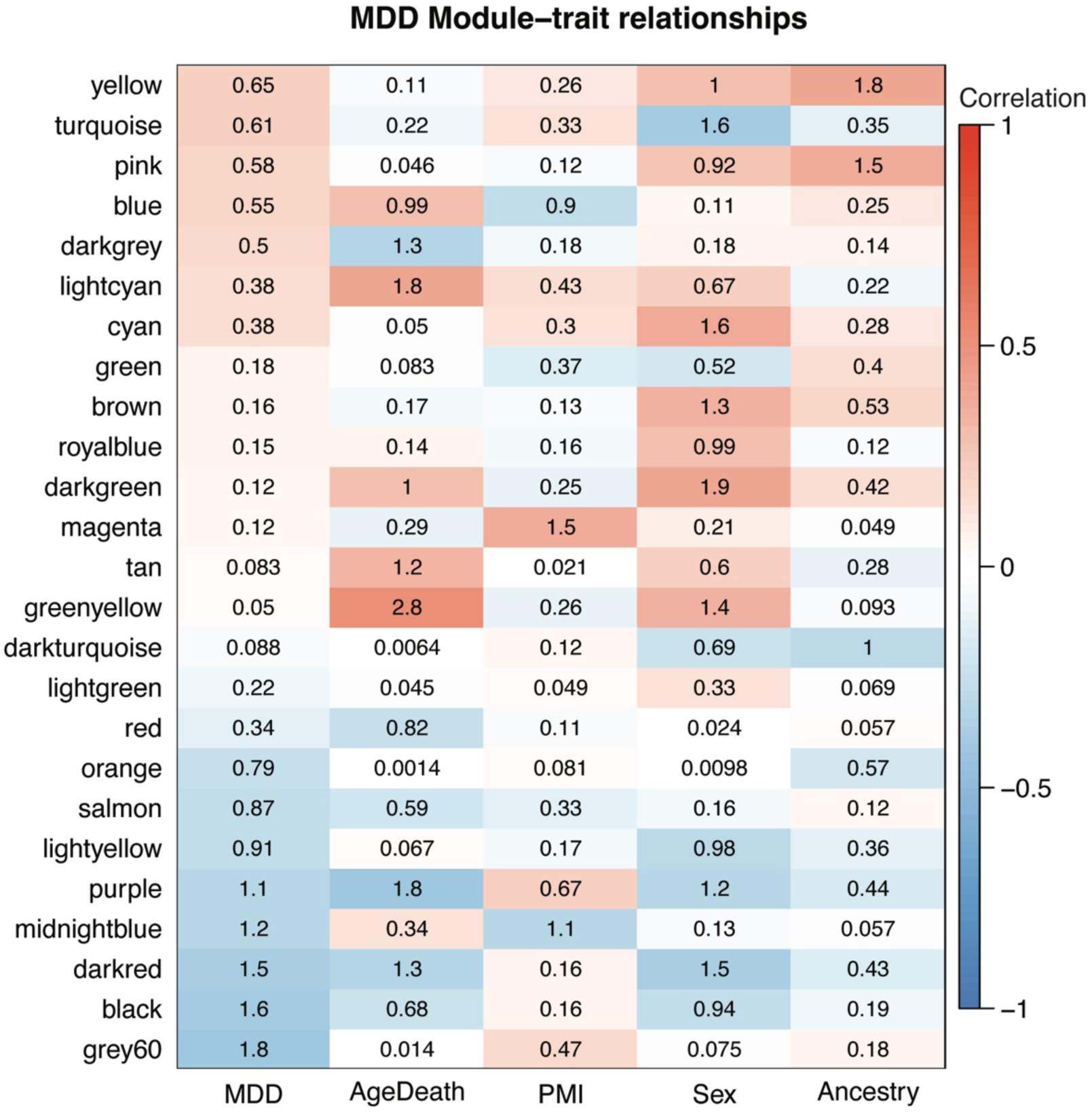
Protein module-trait association for MDD. Color in each cell reflects to correlation between module eigenprotein and PrimaryDx, while the number represents -log_10_(*P*-value) of that correlation.

**Extended Data Fig 4.**
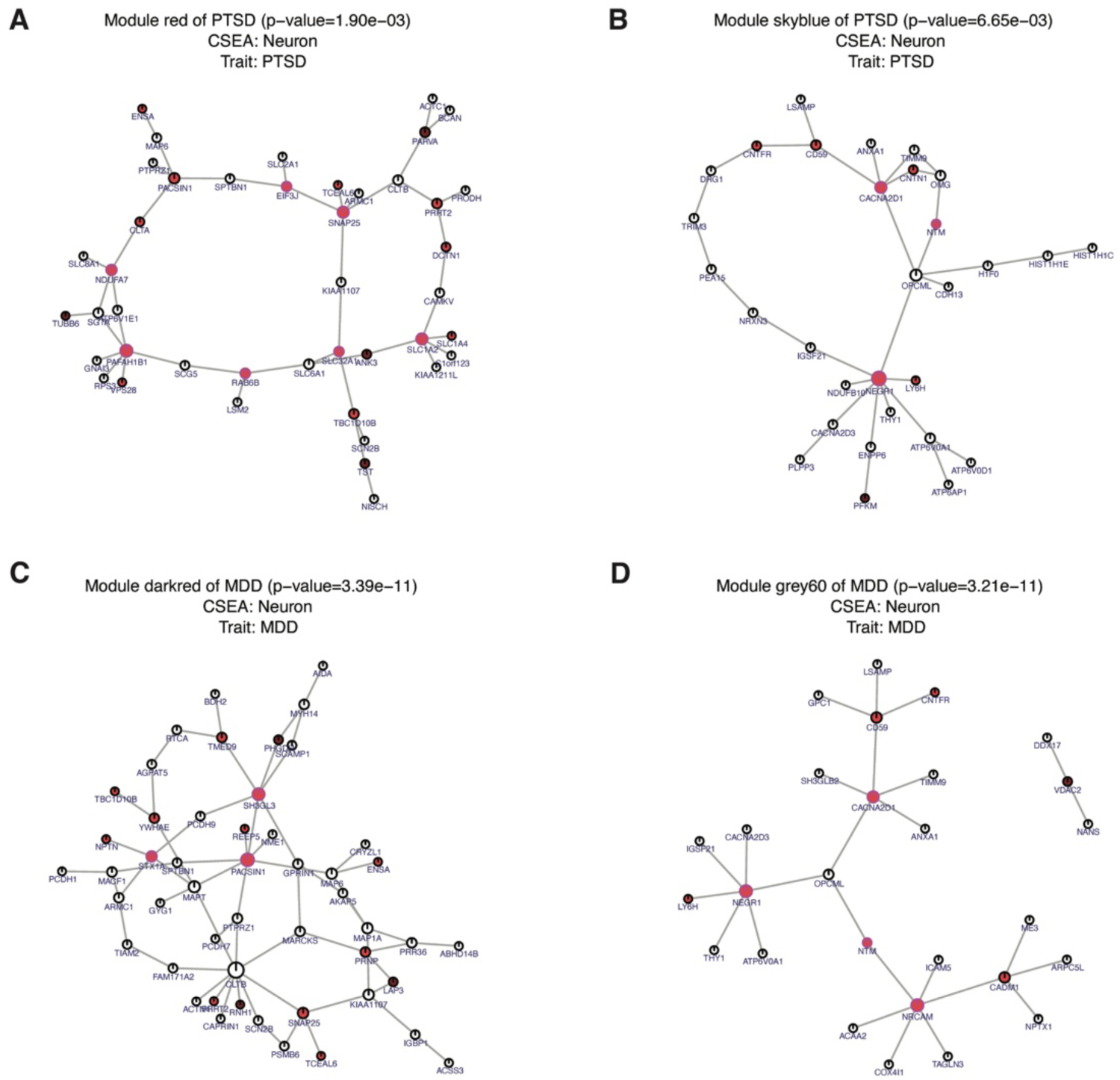
Network and key driver analysis of protein modules. KDA plots for PTSD module *red* (**A**), PTSD module *skyblue* (**B**), MDD module *darkred* (**C**), and MDD module *grey60* (**D**). Key drivers are colored in red and other nodes are in black. Connections indicate significant associations between two proteins. Node sizes measures their total connectivity.

**Extended Data Fig 5.**
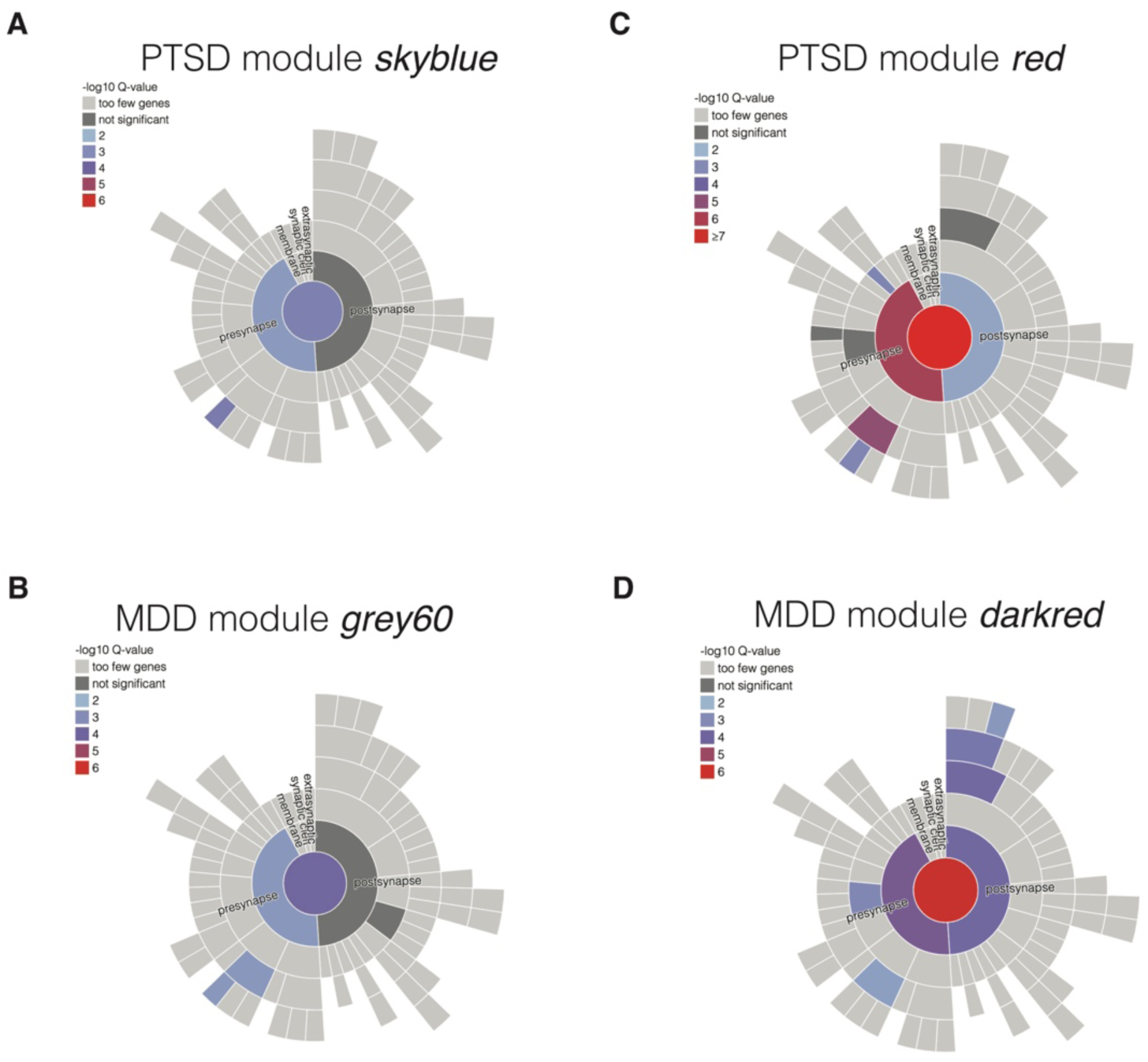
Synaptic function enrichment analysis of protein modules. SynGO plots for (**A**) PTSD module *skyblue*, (**B**) PTSD module *red*, (**C**) MDD module *grey60*, (**D**) MDD module *darkred*. Color legends indicate level of enrichment. Mode SynGO results and statistics are included in **Supp Table 2**.

**Extended Data Fig 6.**
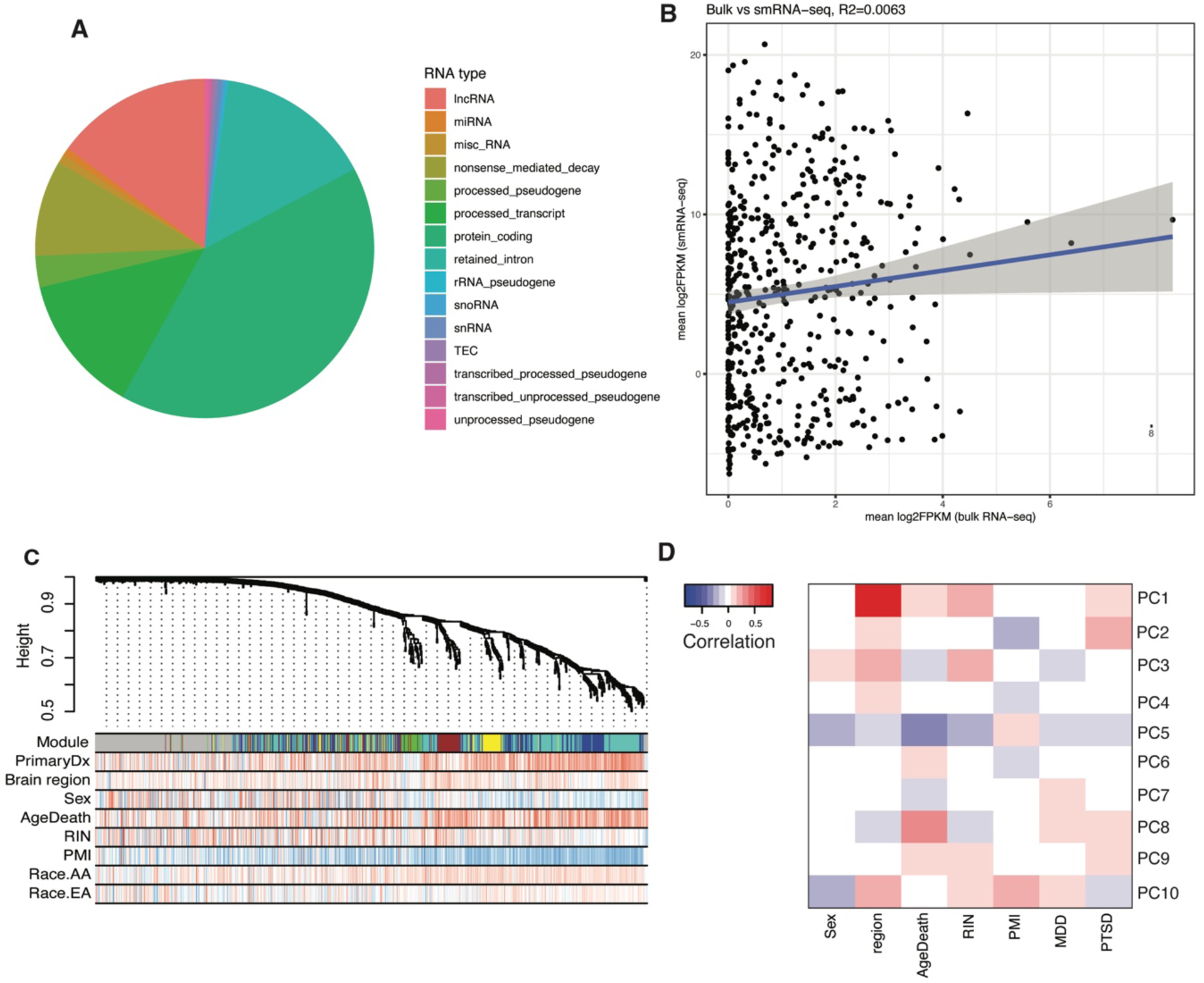
Quality control and cluster analysis of miRNA samples. (**A**) smRNA-seq decomposition of RNA types. (**B**) Comparison between bulk vs smRNA-seq. Bulk RNA-seq and small RNA-seq share low levels of expression of miRNAs (*R*^2^=0.0063). (**C**) miRNA hierarchical clustering dendrogram. (**D**) PCA of miRNAs samples shows brain region is the most significant trait (*P* < 2.2 × 10^-16^ and cor = 0.82) compared to other demographic features (sex, age, RIN, PMI) and diagnosis.

**Extended Data Fig 7.**
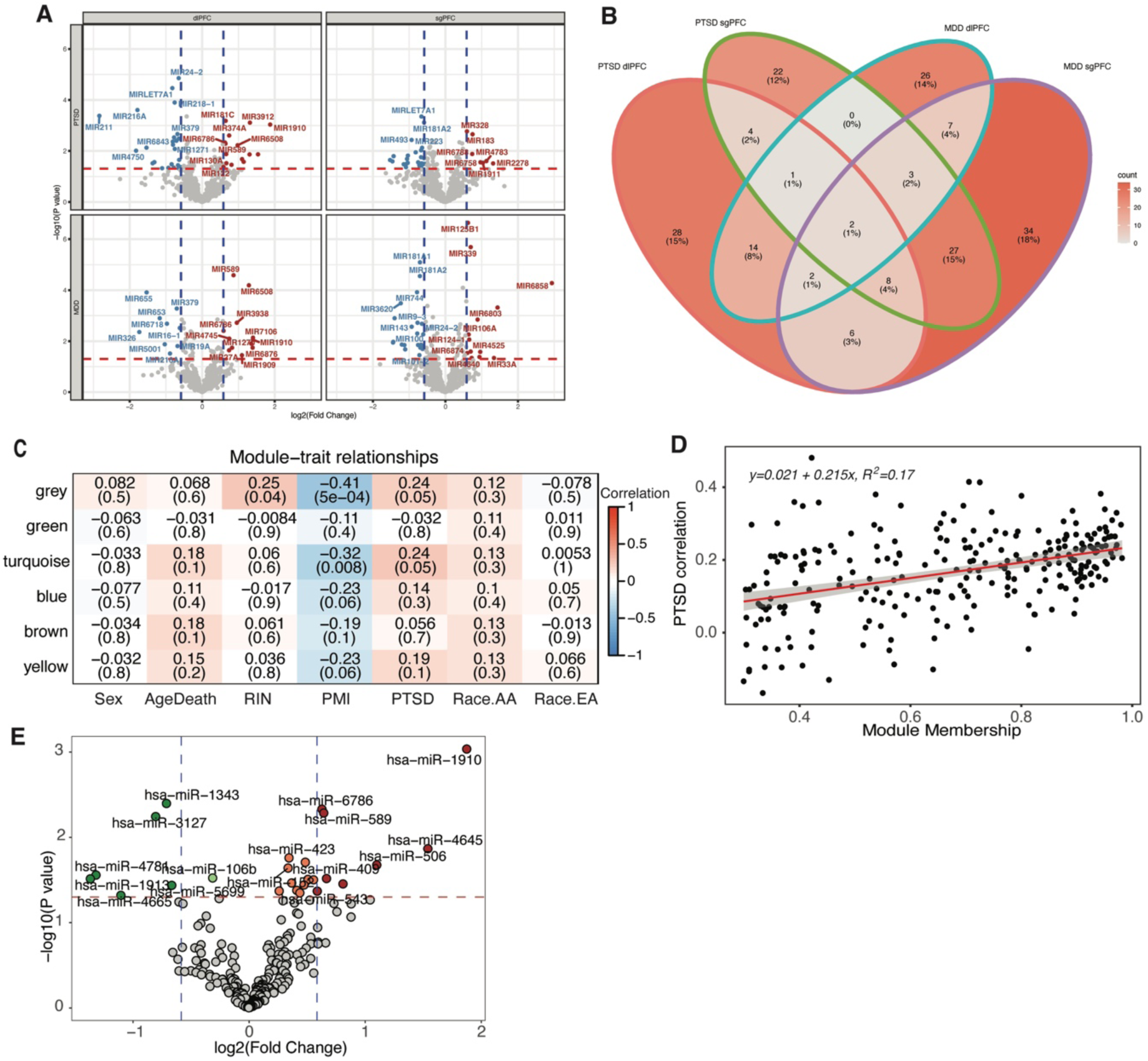
Differential expression and network analysis of miRNA samples. (**A**) Region-specific miRNA volcano plots. Vertical blue dashed lines indicate log fold change threshold of log_2_FC = ±log_2_(1.5). Horizontal red dashed lines indicate significant threshold *P* = 0.05. (**B**) Venn diagram showing overlap of DE miRNAs defined by nominal *P* < 0.05. (**C**) miRNA WGCNA modules. Colors and numbers (out of parentheses) of cells in the heatmap indicate the correlation between the expression module eigengene and trait, while numbers inside parentheses are *P* values of the correlation. (**D**) miRNA module *turquoise* associated with PTSD with an *R*^2^ = 0.17 between the module membership and gene-PTSD diagnosis correlation. (**E**) differential expression of miRNA module *turquoise*. Vertical dashed lines indicate log fold change threshold of log_2_FC = ±log_2_(1.5). Horizontal red dashed lines indicate significance threshold *P* = 0.05. DE miRNAs are marked on the plot as orange (fold change > 1 and *P* < 0.05) and red (fold change > 1.5 and *P* < 0.05), indicating upregulated miRNAs, and green (fold change < 1 and *P* < 0.05) and darkgreen (fold change < 1/1.5 and *P* < 0.05), indicating down regulated ones.

**Extended Data Fig 8.**
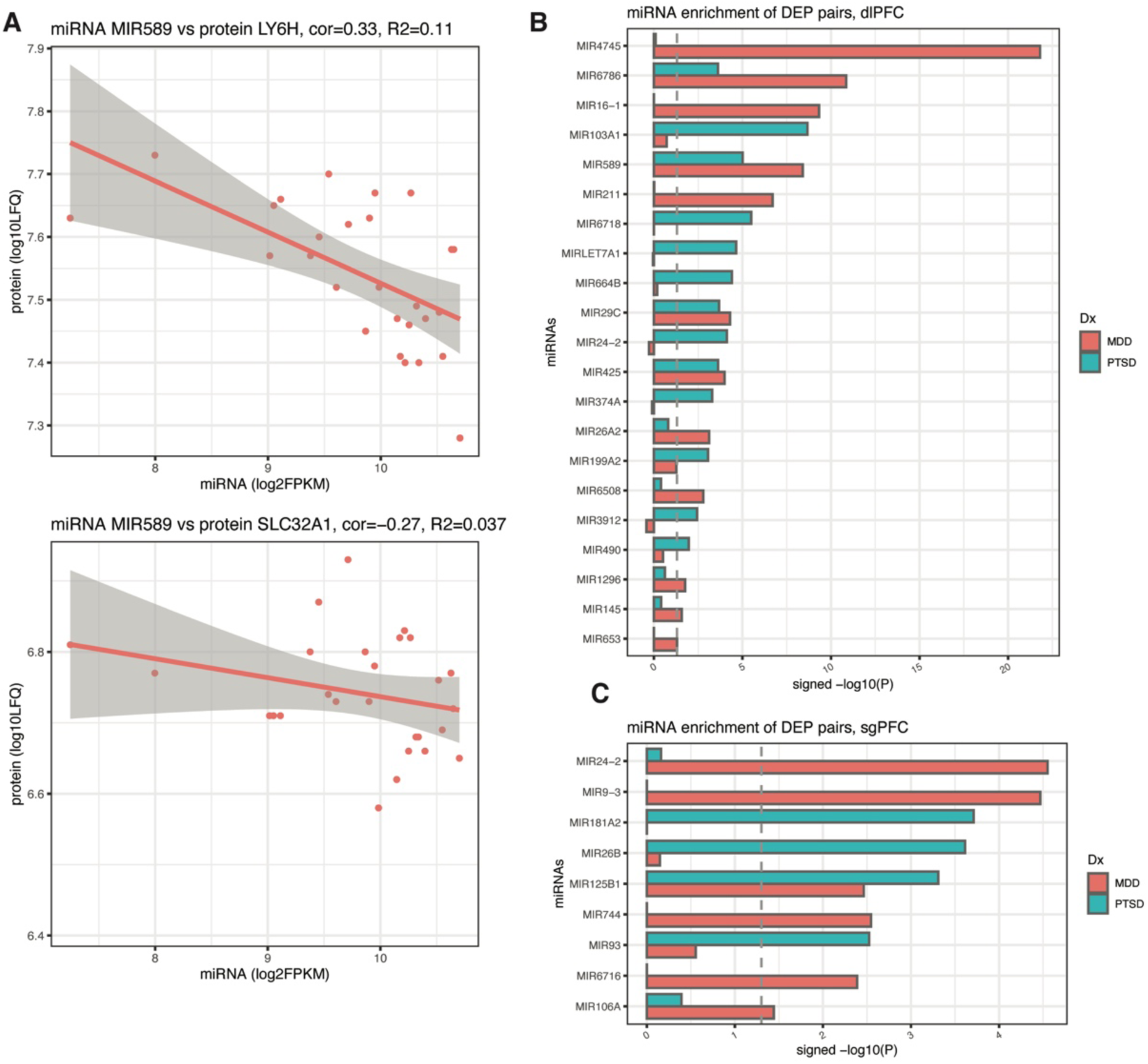
miRNA-protein enrichment analysis. (**A**) Sample correlation between miR589 and protein LY6H (cor = −0.33 and *R*^2^ = 0.11 and protein SLC32A1 (cor = −0.27 and *R*^2^ = 0.037). (**B,C**) List of miRNA-DEP enrichment scores for MDD (red) and PTSD (blue) in dlPFC (**B**) and sgPFC (**C**). Vertical dashed line indicates significant threshold of *P* = 0.05.

**Extended Data Fig 9.**
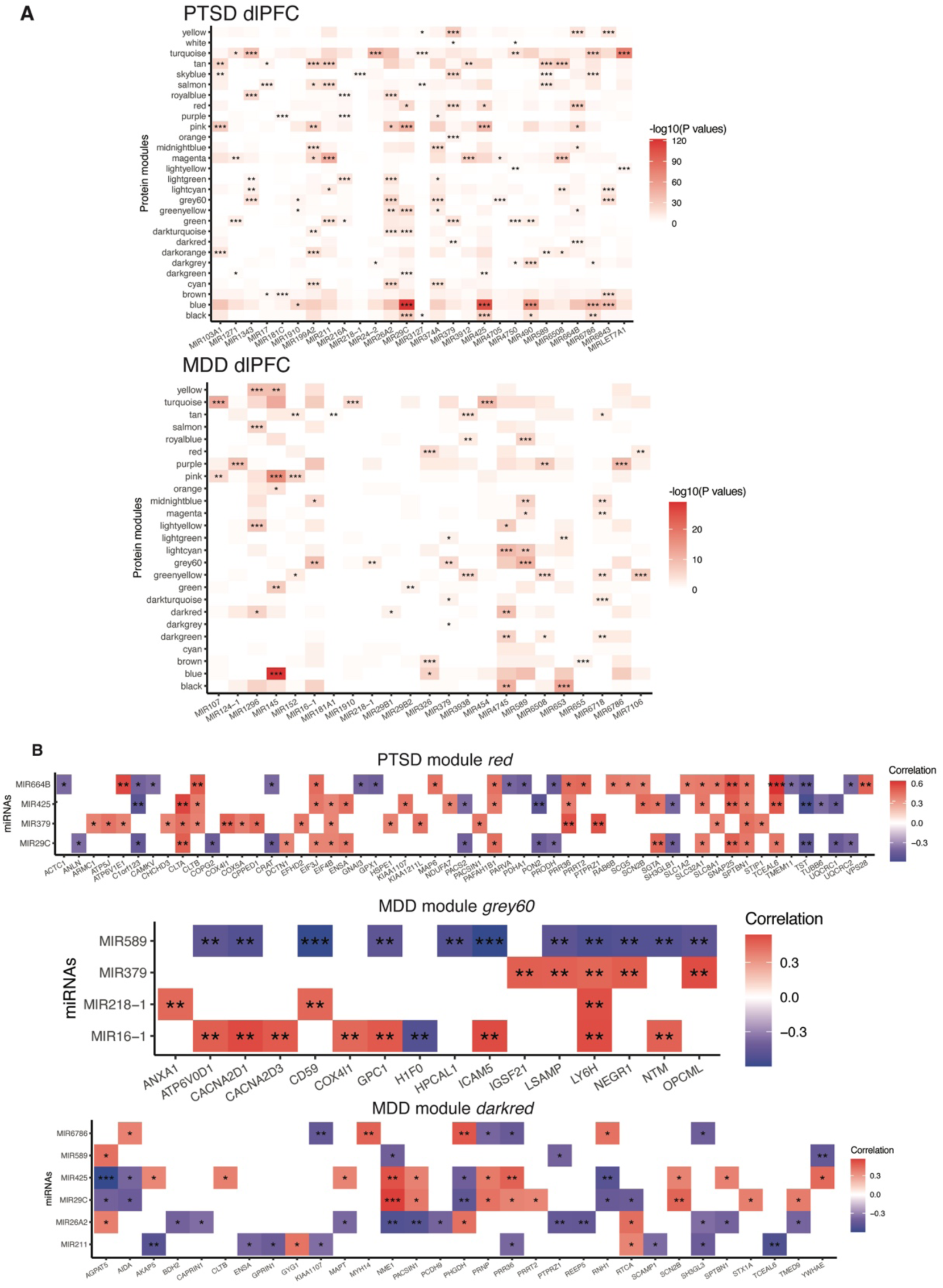
miRNA-protein module enrichment analysis. (**A**) miRNA-module enrichment results for PTSD (top) and MDD (bottom) in DLPFC. Colors of the heatmap indicate the significance levels that are the -log_10_(*P* values) of Fisher’s exact test enrichment scores. (**B**) miRNA-module correlation in PTSD module *red* (top), MDD modules *grey60* (middle) and *darkred* (bottom). Colors in heatmap indicate the correlation levels between miRNAs and proteins. Plotted are miRNA-protein pairs with *P* value < 0.05.

## References

1. Kessler, R. C. & Wang, P. S. The Descriptive Epidemiology of Commonly Occurring Mental Disorders in the United States. Annu. Rev. Public Health 29, 115–129 (2008).

2. Shalev, A., Liberzon, I. & Marmar, C. Post-Traumatic Stress Disorder. http://dx.doi.org/10.1056/NEJMra1612499 https://www.nejm.org/doi/10.1056/NEJMra1612499 (2017) doi:10.1056/NEJMra1612499.

3. Fenster, R. J., Lebois, L. A. M., Ressler, K. J. & Suh, J. Brain circuit dysfunction in post-traumatic stress disorder: from mouse to man. Nat. Rev. Neurosci. 19, 535–551 (2018).

4. Blanchard, E. B., Jones-Alexander, J., Buckley, T. C. & Forneris, C. A. Psychometric properties of the PTSD Checklist (PCL). Behav. Res. Ther. 34, 669–673 (1996).

5. Hankin, C. S., Spiro, A., Miller, D. R. & Kazis, L. Mental Disorders and Mental Health Treatment Among U.S. Department of Veterans Affairs Outpatients: The Veterans Health Study. Am. J. Psychiatry 156, 1924–1930 (1999).

6. Keane, T. M., Taylor, K. L. & Penk, W. E. Differentiating Post-Traumatic Stress Disorder (PTSD) from Major Depression (MDD) and Generalized Anxiety Disorder (GAD). J. Anxiety Disord. 11, 317–328 (1997).

7. Post, L. M., Zoellner, L. A., Youngstrom, E. & Feeny, N. C. Understanding the relationship between co-occurring PTSD and MDD: Symptom severity and affect. J. Anxiety Disord. 25, 1123–1130 (2011).

8. Duncan, L. E. et al. Largest GWAS of PTSD (N=20 070) yields genetic overlap with schizophrenia and sex differences in heritability. Mol. Psychiatry 23, 666–673 (2018).

9. Koenen, K. C. et al. A high risk twin study of combat-related PTSD comorbidity. Twin Res. Off. J. Int. Soc. Twin Stud. 6, 218–226 (2003).

10. Koenen, K. C. et al. A twin registry study of the relationship between posttraumatic stress disorder and nicotine dependence in men. Arch. Gen. Psychiatry 62, 1258– 1265 (2005).

11. Nievergelt, C. M. et al. International meta-analysis of PTSD genome-wide association studies identifies sex- and ancestry-specific genetic risk loci. Nat. Commun. 10, 4558 (2019).

12. Stein, M. B. et al. Genome-wide association analyses of post-traumatic stress disorder and its symptom subdomains in the Million Veteran Program. Nat. Genet. 53, 174–184 (2021).

13. Gelernter, J. et al. Genome-wide association study of post-traumatic stress disorder reexperiencing symptoms in >165,000 US veterans. Nat. Neurosci. 22, 1394–1401 (2019).

14. Girgenti, M. J. et al. Transcriptomic organization of the human brain in post-traumatic stress disorder. Nat. Neurosci. 24, 24–33 (2021).

15. Jaffe, A. E. et al. Decoding Shared Versus Divergent Transcriptomic Signatures Across Cortico-Amygdala Circuitry in PTSD and Depressive Disorders. Am. J. Psychiatry 179, 673–686 (2022).

16. Logue, M. W. et al. Gene expression in the dorsolateral and ventromedial prefrontal cortices implicates immune-related gene networks in PTSD. Neurobiol. Stress 15, 100398 (2021).

17. Dong, P. et al. Population-level variation in enhancer expression identifies disease mechanisms in the human brain. Nat. Genet. 54, 1493–1503 (2022).

18. Kroes, M. C. W., Rugg, M. D., Whalley, M. G. & Brewin, C. R. Structural brain abnormalities common to posttraumatic stress disorder and depression. J. Psychiatry Neurosci. JPN 36, 256–265 (2011).

19. Nees, F., Witt, S. H. & Flor, H. Neurogenetic Approaches to Stress and Fear in Humans as Pathophysiological Mechanisms for Posttraumatic Stress Disorder. Biol. Psychiatry 83, 810–820 (2018).

20. Doerr, A. DIA mass spectrometry. Nat. Methods 12, 35–35 (2015).

21. Langfelder, P. & Horvath, S. WGCNA: an R package for weighted correlation network analysis. BMC Bioinformatics 9, 559 (2008).

22. Xu, X., Wells, A. B., O’Brien, D. R., Nehorai, A. & Dougherty, J. D. Cell Type-Specific Expression Analysis to Identify Putative Cellular Mechanisms for Neurogenetic Disorders. J. Neurosci. 34, 1420–1431 (2014).

23. Koopmans, F. et al. SynGO: An Evidence-Based, Expert-Curated Knowledge Base for the Synapse. Neuron 103, 217–234.e4 (2019).

24. Mahadevan, V. et al. Native KCC2 interactome reveals PACSIN1 as a critical regulator of synaptic inhibition. eLife 6, e28270 (2017).

25. Pérez-Otaño, I. et al. Endocytosis and synaptic removal of NR3A-containing NMDA receptors by PACSIN1/syndapin1. Nat. Neurosci. 9, 611–621 (2006).

26. Ma, D. et al. Antipsychotic Treatment Alters Protein Expression Associated with Presynaptic Function and Nervous System Development in Rat Frontal Cortex. J. Proteome Res. 8, 3284–3297 (2009).

27. Zhang, F. et al. Genetic evidence suggests posttraumatic stress disorder as a subtype of major depressive disorder. J. Clin. Invest. 132, e145942 (2022).

28. Krämer, A., Green, J., Pollard, J. & Tugendreich, S. Causal analysis approaches in Ingenuity Pathway Analysis. Bioinforma. Oxf. Engl. 30, 523–530 (2014).

29. Kasckow, J. W., Baker, D. & Geracioti, T. D. Corticotropin-releasing hormone in depression and post-traumatic stress disorder. Peptides 22, 845–851 (2001).

30. Gandal, M. J. et al. Shared molecular neuropathology across major psychiatric disorders parallels polygenic overlap. Science 359, 693–697 (2018).

31. Kunkle, B. W. et al. Genetic meta-analysis of diagnosed Alzheimer’s disease identifies new risk loci and implicates Aβ, tau, immunity and lipid processing. Nat. Genet. 51, 414–430 (2019).

32. Grove, J. et al. Identification of common genetic risk variants for autism spectrum disorder. Nat. Genet. 51, 431–444 (2019).

33. Stahl, E. A. et al. Genome-wide association study identifies 30 loci associated with bipolar disorder. Nat. Genet. 51, 793–803 (2019).

34. Pardiñas, A. F. et al. Common schizophrenia alleles are enriched in mutation-intolerant genes and in regions under strong background selection. Nat. Genet. 50, 381–389 (2018).

35. Howard, D. M. et al. Genome-wide meta-analysis of depression identifies 102 independent variants and highlights the importance of the prefrontal brain regions. Nat. Neurosci. 22, 343–352 (2019).

36. Hu, Y. et al. A statistical framework for cross-tissue transcriptome-wide association analysis. Nat. Genet. 51, 568–576 (2019).

37. GTEx Consortium. Human genomics. The Genotype-Tissue Expression (GTEx) pilot analysis: multitissue gene regulation in humans. Science 348, 648–660 (2015).

38. Zhang, Y. et al. Differential exosomal microRNA profile in the serum of a patient with depression. Eur. J. Psychiatry 32, 105–112 (2018).

39. Van der Auwera, S. et al. Association of childhood traumatization and neuropsychiatric outcomes with altered plasma micro RNA-levels. Neuropsychopharmacology 44, 2030–2037 (2019).

40. Żurawek, D. & Turecki, G. The miRNome of Depression. Int. J. Mol. Sci. 22, 11312 (2021).

41. Agarwal, V., Bell, G. W., Nam, J.-W. & Bartel, D. P. Predicting effective microRNA target sites in mammalian mRNAs. eLife 4, (2015).

42. McGeary, S. E. et al. The biochemical basis of microRNA targeting efficacy. Science 366, eaav1741 (2019).

43. Griffiths-Jones, S. The microRNA Registry. Nucleic Acids Res. 32, D109–D111 (2004).

44. Griffiths-Jones, S., Grocock, R. J., van Dongen, S., Bateman, A. & Enright, A. J. miRBase: microRNA sequences, targets and gene nomenclature. Nucleic Acids Res. 34, D140–D144 (2006).

45. Griffiths-Jones, S., Saini, H. K., van Dongen, S. & Enright, A. J. miRBase: tools for microRNA genomics. Nucleic Acids Res. 36, D154–D158 (2008).

46. Kozomara, A. & Griffiths-Jones, S. miRBase: integrating microRNA annotation and deep-sequencing data. Nucleic Acids Res. 39, D152–D157 (2011).

47. Kozomara, A. & Griffiths-Jones, S. miRBase: annotating high confidence microRNAs using deep sequencing data. Nucleic Acids Res. 42, D68–D73 (2014).

48. Kozomara, A., Birgaoanu, M. & Griffiths-Jones, S. miRBase: from microRNA sequences to function. Nucleic Acids Res. 47, D155–D162 (2019).

49. Girgenti, M. J. & Duman, R. S. Transcriptome Alterations in Posttraumatic Stress Disorder. Biol. Psychiatry 83, 840–848 (2018).

50. Alexandra Kredlow, M., Fenster, R. J., Laurent, E. S., Ressler, K. J. & Phelps, E. A. Prefrontal cortex, amygdala, and threat processing: implications for PTSD. Neuropsychopharmacology 47, 247–259 (2022).

51. Bremner, J. D. et al. Neural Correlates of Memories of Childhood Sexual Abuse in Women With and Without Posttraumatic Stress Disorder. Am. J. Psychiatry 156, 1787–1795 (1999).

52. Liberzon, I. et al. Brain activation in PTSD in response to trauma-related stimuli. Biol. Psychiatry 45, 817–826 (1999).

53. Shin, L. M. Amygdala, Medial Prefrontal Cortex, and Hippocampal Function in PTSD. Ann. N. Y. Acad. Sci. 1071, 67–79 (2006).

54. Shin, L. M. et al. Regional Cerebral Blood Flow in the Amygdala and Medial PrefrontalCortex During Traumatic Imagery in Male and Female Vietnam Veterans With PTSD. Arch. Gen. Psychiatry 61, 168 (2004).

55. Li, H. et al. Functional annotation of the human PTSD methylome identifies tissue-specific epigenetic variation across subcortical brain regions. Preprint at https://doi.org/10.1101/2023.04.18.23288704 (2023).

56. Tomioka, N. H. et al. Elfn1 recruits presynaptic mGluR7 in trans and its loss results in seizures. Nat. Commun. 5, 4501 (2014).

57. Liu, Y. et al. PACSIN1, a Tau-interacting Protein, Regulates Axonal Elongation and Branching by Facilitating Microtubule Instability. J. Biol. Chem. 287, 39911–39924 (2012).

58. Regan, P. et al. Regulation of Synapse Weakening through Interactions of the Microtubule Associated Protein Tau with PACSIN1. J. Neurosci. Off. J. Soc. Neurosci. 41, 7162–7170 (2021).

59. Puddifoot, C. A., Wu, M., Sung, R.-J. & Joiner, W. J. Ly6h Regulates Trafficking of Alpha7 Nicotinic Acetylcholine Receptors and Nicotine-Induced Potentiation of Glutamatergic Signaling. J. Neurosci. 35, 3420–3430 (2015).

60. Panichareon, B., Nakayama, K., Thurakitwannakarn, W., Iwamoto, S. & Sukhumsirichart, W. OPCML Gene as a Schizophrenia Susceptibility Locus in Thai Population. J. Mol. Neurosci. 46, 373–377 (2012).

61. Schol-Gelok, S. et al. A Genome-Wide Screen for Depression in Two Independent Dutch Populations. Biol. Psychiatry 68, 187–196 (2010).

62. Pischedda, F. & Piccoli, G. The IgLON Family Member Negr1 Promotes Neuronal Arborization Acting as Soluble Factor via FGFR2. Front. Mol. Neurosci. 8, (2016).

63. Brugger, S. W., Gardner, M. C., Beales, J. T., Briggs, F. & Davis, M. F. Depression in multiple sclerosis patients associated with risk variant near NEGR1. Mult. Scler. Relat. Disord. 46, 102537 (2020).

64. Levey, D. F. et al. Bi-ancestral depression GWAS in the Million Veteran Program and meta-analysis in >1.2 million individuals highlight new therapeutic directions. Nat. Neurosci. 24, 954–963 (2021).

65. Wang, X. et al. Integrating genome-wide association study and expression quantitative trait loci data identifies NEGR1 as a causal risk gene of major depression disorder. J. Affect. Disord. 265, 679–686 (2020).

66. Wray, N. R. et al. Genome-wide association analyses identify 44 risk variants and refine the genetic architecture of major depression. Nat. Genet. 50, 668–681 (2018).

67. Noh, K. et al. Negr1 controls adult hippocampal neurogenesis and affective behaviors. Mol. Psychiatry 24, 1189–1205 (2019).

68. Gamero-Villarroel, C. et al. Impact of NEGR1 genetic variability on psychological traits of patients with eating disorders. Pharmacogenomics J. 15, 278–283 (2015).

69. Karis, K. et al. Altered Expression Profile of IgLON Family of Neural Cell Adhesion Molecules in the Dorsolateral Prefrontal Cortex of Schizophrenic Patients. Front. Mol. Neurosci. 11, 8 (2018).

70. Zhang, T. et al. Voltage-gated calcium channel activity and complex related genes and schizophrenia: A systematic investigation based on Han Chinese population. J. Psychiatr. Res. 106, 99–105 (2018).

71. Lutz, M. W., Luo, S., Williamson, D. E. & Chiba-Falek, O. Shared genetic etiology underlying late-onset Alzheimer’s disease and posttraumatic stress syndrome. Alzheimers Dement. J. Alzheimers Assoc. 16, 1280–1292 (2020).

72. Flatt, J. D., Gilsanz, P., Quesenberry, C. P., Albers, K. B. & Whitmer, R. A. Post-traumatic stress disorder and risk of dementia among members of a health care delivery system. Alzheimers Dement. 14, 28–34 (2018).

73. Logue, M. W. et al. Alzheimer’s disease and related dementias among aging veterans: Examining gene-by-environment interactions with post-traumatic stress disorder and traumatic brain injury. Alzheimers Dement. alz.12870 (2022) doi:10.1002/alz.12870.

74. Gugliandolo, A. et al. MicroRNAs Modulate the Pathogenesis of Alzheimer’s Disease: An In Silico Analysis in the Human Brain. Genes 11, 983 (2020).

75. Lu, L., Dai, W.-Z., Zhu, X.-C. & Ma, T. Analysis of Serum miRNAs in Alzheimer’s Disease. Am. J. Alzheimers Dis. Dementias® 36, 15333175211021712 (2021).

76. Jiang, Q. et al. miR-24 protects against ischemia-induced brain damage in rats via regulating microglia polarization by targeting Clcn3. Neurosci. Lett. 759, 135998 (2021).

77. Villela, D. et al. Differential DNA Methylation of MicroRNA Genes in Temporal Cortex from Alzheimer’s Disease Individuals. Neural Plast. 2016, e2584940 (2016).

78. Wingo, T. S. et al. Brain microRNAs associated with late-life depressive symptoms are also associated with cognitive trajectory and dementia. Npj Genomic Med. 5, 6 (2020).

79. Margolin, A. A. et al. ARACNE: An Algorithm for the Reconstruction of Gene Regulatory Networks in a Mammalian Cellular Context. BMC Bioinformatics 7, S7 (2006).

